# The impact of the COVID-19 pandemic on Antipsychotic Prescribing in individuals with autism, dementia, learning disability, serious mental illness or living in a care home: A federated analysis of 59 million patients’ primary care records in situ using OpenSAFELY

**DOI:** 10.1101/2023.01.05.23284214

**Authors:** The OpenSAFELY Collaborative:, Orla Macdonald, Amelia Green, Alex Walker, Richard Croker, Helen Curtis, Andrew Brown, Ben Butler-Cole, Colm Andrews, Caroline Morton, Dave Evans, Peter Inglesby, Iain Dillingham, Jon Massey, Louis Fisher, Seb Bacon, Simon Davy, Tom Ward, Will Hulme, Jess Morley, Amir Mehrkar, Chris Bates, Jonathan Cockburn, John Parry, Frank Hester, Sam Harper, Shaun O’Hanlon, Alex Eavis, Richard Jarvis, Dima Avramov, Ian Wood, Nasreen Parkes, Ben Goldacre, Brian MacKenna

## Abstract

**Background:** The COVID-19 pandemic significantly affected health and social care services. We aimed to explore whether this impacted the prescribing rates of antipsychotics within at-risk populations.

**Methods:** With the approval of NHS England, we completed a retrospective cohort study, using the OpenSAFELY platform to explore primary care data of 59 million patients. We identified patients in five at-risk groups: autism, dementia, learning disability, serious mental illness and care home residents. We then calculated the monthly prevalence of antipsychotic prescribing in the population, as well as the incidence of new prescriptions in each month over the study period (Jan 2019-Dec 2021).

**Results:** The average monthly rate of antipsychotic prescribing increased in dementia from 82.75 patients prescribed an antipsychotic per 1000 patients (95% CI 82.30-83.19) in Q1 2019 to 90.1 (95% CI 89.68-90.60) in Q4 2021 and from 154.61 (95% CI 153.79-155.43) in Q1 2019 to 166.95 (95% CI 166.23-167.67) in Q4 2021 in care homes. There were notable spikes in the rate of new prescriptions issued to patients with dementia and in care homes. In learning disability and autism groups, the average monthly rate of prescribing per 1000 decreased from 122.97 (95% CI 122.29-123.66) in Q1 2019 to 119.29 (95% CI 118.68-119.91) in Q4 2021, and from 54.91 (95% CI 54.52-55.29) in Q1 2019 to 51.04 (95% CI 50.74-51.35) in Q4 2021 respectively.

**Conclusions:** During each of the lockdowns in 2020, we observed a significant spike in antipsychotic prescribing in the dementia and care home groups. We have shown that these peaks are likely due to prescribing of antipsychotics for palliative care purposes and may have been linked to pre-emptive prescribing, when on-site medical visits would have been restricted. Over the study period, we observed gradual increases in antipsychotic use in patients with dementia and in care homes and a decrease in their use in patients with learning disability or autism.

## BACKGROUND

In March 2020, the UK went into its first national lockdown following the outbreak of the SARS-CoV-2 pandemic. Subsequent literature has described the negative psychological impact of these lockdowns on “at risk” groups of patients, such as those with dementia [1,2], learning disability [3] and autism [4,5], including increases in agitation, anxiety, depressive symptoms and negative behaviours such as aggressive and self harming behaviours, which are sometimes treated with antipsychotics.

Prior to the pandemic the NHS in England had recognised that prescribing of antipsychotics was excessive in certain at-risk populations including in patients with dementia, learning disability, autism and those in care homes. In response, national campaigns, such as the National Dementia Strategy (2009) and STOMP (2016) (Stopping OverMedication of People with a learning disability, autism or both), were funded to reduce the overprescribing of antipsychotics in these groups [6,7]. NICE states that in patients with learning disability or dementia, antipsychotics should only be used to manage behaviour where patients are severely distressed or at risk of harming themselves or others [8,9] and advocates the use of non-pharmacological strategies, including structured activities and positive social support as a preferred approach to managing challenging behaviours. However, many of the social and charitable structures that supported people with disabilities were substantially affected during the pandemic [6]. We were interested to know if the combination of negative impacts of the pandemic led to an increase in antipsychotic use within these at-risk populations.

OpenSAFELY is a new secure analytics platform for electronic patient records built by our group on behalf of NHS England to deliver urgent academic [10,11,12] and operational research during the pandemic [13,14,15]. One of the aims of this research platform is to assess the effect of the pandemic on indirect health related outcomes; such as the impact of the dramatic change in services on at-risk populations. OpenSAFELY analyses can currently run across all patients’ full raw pseudonymised primary care records at 99% of English general practices, with patient-level linkage to various sources of secondary care data. All code and analysis is shared openly for inspection and re-use.

We therefore set out to use the OpenSAFELY platform to assess the impact of the COVID-19 pandemic on antipsychotic prescribing trends in at-risk populations and to establish if there was a change in prescribing practice.

## METHODS

### Study Design

We conducted a retrospective population-based cohort study using general practice primary care electronic health record (EHR) data accessed through OpenSAFELY, currently covering 99% of England’s general practices. Data was included from January 2019 to December 2021 and therefore included 14 months of prescribing prior to the first lockdown in March 2020 and 22 months after this point.

### Data Source

Primary care records for all practices in England managed by the GP software providers TPP and EMIS are available in OpenSAFELY, a data analytics platform created by our team on behalf of NHS England to address urgent COVID-19 research questions (https://opensafely.org). OpenSAFELY provides a secure software interface allowing the analysis of pseudonymized primary care patient records from England in near real-time within the EHR vendor’s highly secure data centre, avoiding the need for large volumes of potentially disclosive pseudonymized patient data to be transferred off-site. This, in addition to other technical and organisational controls, minimises any risk of re-identification. The TPP dataset analysed within OpenSAFELY (OpenSAFELY-TPP) is based on 24.8 million people currently registered with GP surgeries using TPP SystmOne software; the EMIS dataset analysed within OpenSAFELY (OpenSAFELY-EMIS) is based on 35.2 million people currently registered with GP surgeries using EMIS. It includes pseudonymized data such as coded diagnoses, medications and physiological parameters. No free text data are included. Further details on our information governance can be found in the information governance and ethics section below.

### Study population

We included all individuals who were alive and registered at an OpenSAFELY-TPP or OpenSAFELY-EMIS practice each month, across the study period.

### At-Risk groups

Five at-risk groups were identified consisting of individuals who had a recorded history of a diagnosis of learning disability, autism, dementia, serious mental illness or a history of being a resident in a nursing or residential care home according to their GP record. These groups were non-exclusive (e.g., patients in a care home and with dementia were counted in both groups). For the care home group, we included all those coded by GPs as living in residential accommodation, however this is not a comprehensive list of all patients who live in supported accommodation [16]. For our dementia cohort we excluded any patient under the age of 35, where a diagnosis of dementia is extremely rare. Further details can be found under the codelists and implementation section below.

### Antipsychotic medications

Prescriptions for antipsychotic medications were defined using NHS Dictionary of Medicines and Devices codes (codelist details can be found in the appendices). We categorised antipsychotic medication in terms of whether they were first or second generation, or long-acting injectable and depot antipsychotics, and those that are routinely used for other indications, such as prochlorperazine which is used extensively for nausea and vomiting (Table S3). We excluded antipsychotics which are exclusively used in palliative care, such as the injectable forms of levomepromazine. Over the study period, only the most recently available specified code for a prescription of an antipsychotic medication within each month was included.

### Population characteristics

To summarise the population we counted patients registered at the end of the period (1^st^ October and 31^st^ December 2021) and the number of whom received one or more prescriptions for an antipsychotic during this period, and we described the demographics of this subgroup compared to the total population. This was repeated for each at-risk group.

We extracted patient demographic information on age (categorised into eight classes), sex, 2019 Index of Multiple Deprivation (IMD) as quintiles, ethnicity (17 categories) and region (7 NHS England regions). Patient demographics were extracted monthly, except ethnicity which (for reduced computational time) was determined once at the end of the study period.

### Trends in antipsychotic prescribing

We counted the number of patients who were issued an antipsychotic prescription each month between January 2019 and December 2021. We calculated this for the population as a whole and for each of the five at-risk groups. From this we calculated both a monthly and quarterly rate (per 1000 eligible population) of antipsychotic prescribing for each group and for the population as a whole. The quarterly rate is preferred because there is more variation at a monthly level and there are expected anomalies with prescribing data in certain months (e.g. February and December) due to reduced GP working hours within those months, however the monthly data was included so that we could show significant changes in prescribing that were relatively short lived.

### Trends in patients newly prescribed antipsychotics

We counted the number of patients who were newly prescribed an antipsychotic (those with no recorded antipsychotic prescription in the 24 months prior) each month between January 2019 and December 2021. We calculated the rate (per 1000 eligible population) of newly initiated patients for the population as a whole and in each at-risk group.

### Sensitivity analysis

We completed a further sensitivity analysis in patients with dementia and in care homes to identify if newly prescribed antipsychotics were being initiated for palliative care purposes. For this analysis, we excluded any patient who had died within 2 weeks of the antipsychotic being initiated (one indicator of palliative care) and any patient who had been prescribed midazolam, a drug commonly used in palliative care, at the same time (+/- 1 day) as the antipsychotic.

### Missing data

Individuals with missing ethnicity, IMD and region were included as “Unknown”. Individuals were excluded if they had an unknown date of birth or unknown sex.

### Statistical methods

Simple descriptive statistics were used to summarise the number of patients issued antipsychotic prescriptions, the number of first antipsychotic prescriptions and the monthly rate of antipsychotic prescribing. Rates, with associated 95% confidence intervals (CIs), were estimated by dividing antipsychotic counts by the total at-risk population and multiplying by 1000. Counts and rates of antipsychotic prescriptions were stratified by each demographic variable, within each of the five at-risk populations.

Charts and results not presented in this manuscript are available online for inspection in the associated GitHub repository [17]. Patient counts of 0-7 are shown as “<8” with remaining counts rounded to the nearest 10 to protect against small number differences.

### Codelists and implementation

Information on all covariates were obtained from primary care records by searching TPP/EMIS records for specific coded data. Detailed information on compilation and sources for every individual codelist is available at https://www.opencodelists.org/ and the lists are available for inspection and re-use by the broader research community. Links to specific codelists used within this study can be found in the appendices. All codelists were verified and reviewed by contributors from within the Datalab team.

### Software and Reproducibility

Data management was performed using the OpenSAFELY software libraries, implemented using Python 3.8, with analysis carried out using both Python and R version 4.0.2. This was an analysis delivered using federated analysis through the OpenSAFELY platform. A federated analysis involves carrying out patient level analysis in multiple secure datasets, then later combining them: codelists and code for data management and data analysis were specified once using the OpenSAFELY tools; then transmitted securely from the OpenSAFELY jobs server to the OpenSAFELY-TPP platform within TPP’s secure environment, and separately to the OpenSAFELY-EMIS platform within EMIS’s secure environment, where they were each executed separately against local patient data; summary results were then reviewed for disclosiveness, released, and combined for the final outputs. All code for the OpenSAFELY platform for data management, analysis and secure code execution is shared for review and re-use under open licences on GitHub: https://github.com/opensafely/antipsychotics-prescribing-during-COVID-19.

### Patient and Public Involvement

This analysis relies on the use of large volumes of patient data. Ensuring patient, professional and public trust is therefore of critical importance. Maintaining trust requires being transparent about the way OpenSAFELY works, and ensuring patient voices are represented in the design of research, analysis of the findings, and considering the implications. For transparency purposes we have developed a public website (https://opensafely.org/) which provides a detailed description of the platform in language suitable for a lay audience; we have participated in two citizen juries exploring public trust in OpenSAFELY [18]; we are currently co-developing an explainer video; we have ‘expert by experience’ patient representation on our OpenSAFELY Oversight Board; we have partnered with Understanding Patient Data to produce lay explainers on the importance of large datasets for research; we have presented at a number of online public engagement events to key communities; and more. To ensure the patient voice is represented, we are working closely with appropriate medical research charities.

## RESULTS

### Population characteristics

Between 1^st^ October and 31^st^ December 2021, 59,968,090 individuals were alive and registered at a TPP or EMIS GP practice. Demographics of this cohort are described in Table 1. Within this total population, 543,220 (<1%) patients were prescribed an antipsychotic. Rates of antipsychotic prescribing increased with increasing age and with increasing levels of deprivation; 29% of individuals prescribed an antipsychotic were in the most deprived quintile compared to 12% in the least deprived. A higher proportion of those issued an antipsychotic were female (57%); the baseline population had a 50:50 ratio of women to men.

**Table 1.** Demographics of cohort, split by antipsychotic use.

| Attribute | Category | Combined figures from TPP and EMIS |  |  |
| --- | --- | --- | --- | --- |
|  |  | Total (%) | Not on anti-psychotic (%) | Taking anti- psychotic (%) |
| **Total** |  | 59,968,090 (100) | 59,424,880 (100) | 543,220 (100) |
| **Age group** | **0-17** | 11,283,265 (19) | 11,278,660 (19) | 4,610 (1) |
|  | **18-24** | 4,867,130 (8) | 4,845,160 (8) | 21,970 (4) |
|  | **25-34** | 8,647,075 (14) | 8,584,350 (14) | 62,730 (12) |
|  | **35-44** | 8,543,285 (14) | 8,462,220 (14) | 81,065 (15) |
|  | **45-54** | 7,766,895 (13) | 7,668,135 (13) | 98,765 (18) |
|  | **55-69** | 10,505,365 (18) | 10,364,955 (17) | 140,410 (26) |
|  | **70-79** | 5,099,270 (9) | 5,031,145 (8) | 68,125 (13) |
|  | **80+** | 3,255,805 (5) | 3,190,255 (5) | 65,545 (12) |
| **Sex** | **Female** | 29,937,970 (50) | 29,627,580 (50) | 310,390 (57) |
|  | **Male** | 30,030,130 (50) | 29,797,300 (50) | 232,825 (43) |
| **IMD** | **1 Most deprived** | 12,178,640 (20) | 12,018,870 (20) | 159,765 (29) |
|  | **2** | 12,376,850 (21) | 12,251,495 (21) | 125,350 (23) |
|  | **3** | 11,938,615 (20) | 11,836,940 (20) | 101,680 (19) |
|  | **4** | 11,372,110 (19) | 11,290,090 (19) | 82,020 (15) |
|  | **5 Least deprived** | 11,217,270 (19) | 11,150,375 (19) | 66,895 (12) |
|  | **Unknown** | 884,615 (1) | 877,115 (1) | 7,505 (1) |
| **Ethnicity** | **African** | 1,061,380 (2) | 1,053,880 (2) | 7,505 (1) |
|  | **Bangladeshi or British Bangladeshi** | 474,600 (1) | 470,015 (1) | 4,585 (1) |
|  | **British or Mixed British** | 26,544,735 (44) | 26,229,235 (44) | 315,495 (58) |
|  | **Caribbean** | 375,610 (1) | 370,040 (1) | 5,570 (1) |
|  | **Chinese** | 485,385 (1) | 484,320 (1) | 1,060 (0) |
|  | **Indian or British Indian** | 1,547,965 (3) | 1,538,495 (3) | 9,460 (2) |
|  | **Irish** | 276,660 (0) | 272,495 (0) | 4,170 (1) |
|  | **Other** | 845,665 (1) | 840,120 (1) | 5,545 (1) |
|  | **Other Asian** | 975,140 (2) | 968,960 (2) | 6,175 (1) |
|  | **Other Black** | 270,610 (0) | 267,710 (0) | 2,895 (1) |
|  | **Other mixed** | 375,265 (1) | 372,765 (1) | 2,500 (0) |
|  | **Other White** | 4,208,045 (7) | 4,189,030 (7) | 19,010 (3) |
|  | **Pakistani or British Pakistani** | 1,206,855 (2) | 1,196,145 (2) | 10,715 (2) |
|  | **White + Asian** | 192,705 (0) | 191,515 (0) | 1,190 (0) |
|  | **White + Black African** | 196,975 (0) | 195,685 (0) | 1,290 (0) |
|  | **White + Black Caribbean** | 213,695 (0) | 211,110 (0) | 2,585 (0) |
|  | **Unknown** | 20,716,810 (35) | 20,573,350 (35) | 143,455 (26) |
| **Region** | **East of England** | 7,075,760 (12) | 7,008,010 (12) | 67,750 (12) |
|  | **London** | 10,123,045 (17) | 10,048,125 (17) | 74,920 (14) |
|  | **Midlands** | 11,268,450 (19) | 11,157,880 (19) | 110,565 (20) |
|  | **North East and Yorkshire** | 7,441,485 (12) | 7,376,235 (12) | 65,250 (12) |
|  | **North West** | 8,959,840 (15) | 8,867,515 (15) | 92,325 (17) |
|  | **South East** | 9,108,570 (15) | 9,028,185 (15) | 80,385 (15) |
|  | **South West** | 5,956,165 (10) | 5,904,495 (10) | 51,670 (10) |
|  | **Unknown** | 34,785 (0) | 34,445 (0) | 345 (0) |

### Trends in antipsychotic prescribing

There was a slight increase in the rate of antipsychotic prescribing (per 1000 patients) over the study period from 8.67 (95%CI 8.66-8.68) in Jan-Mar 2019 to 9.09 (95%CI 9.07-9.10) in Oct-Dec 2021 (Figure 1). There was decrease in the overall rate of new initiation of antipsychotics around the time of the first lockdown; in Q2 of 2020 this figure was 0.794 (95%CI 0.791-0.799) whereas in Q2 2019 it was 0.895 (95%CI 0.891-0.899)(Figure 2)

**Figure 1.**
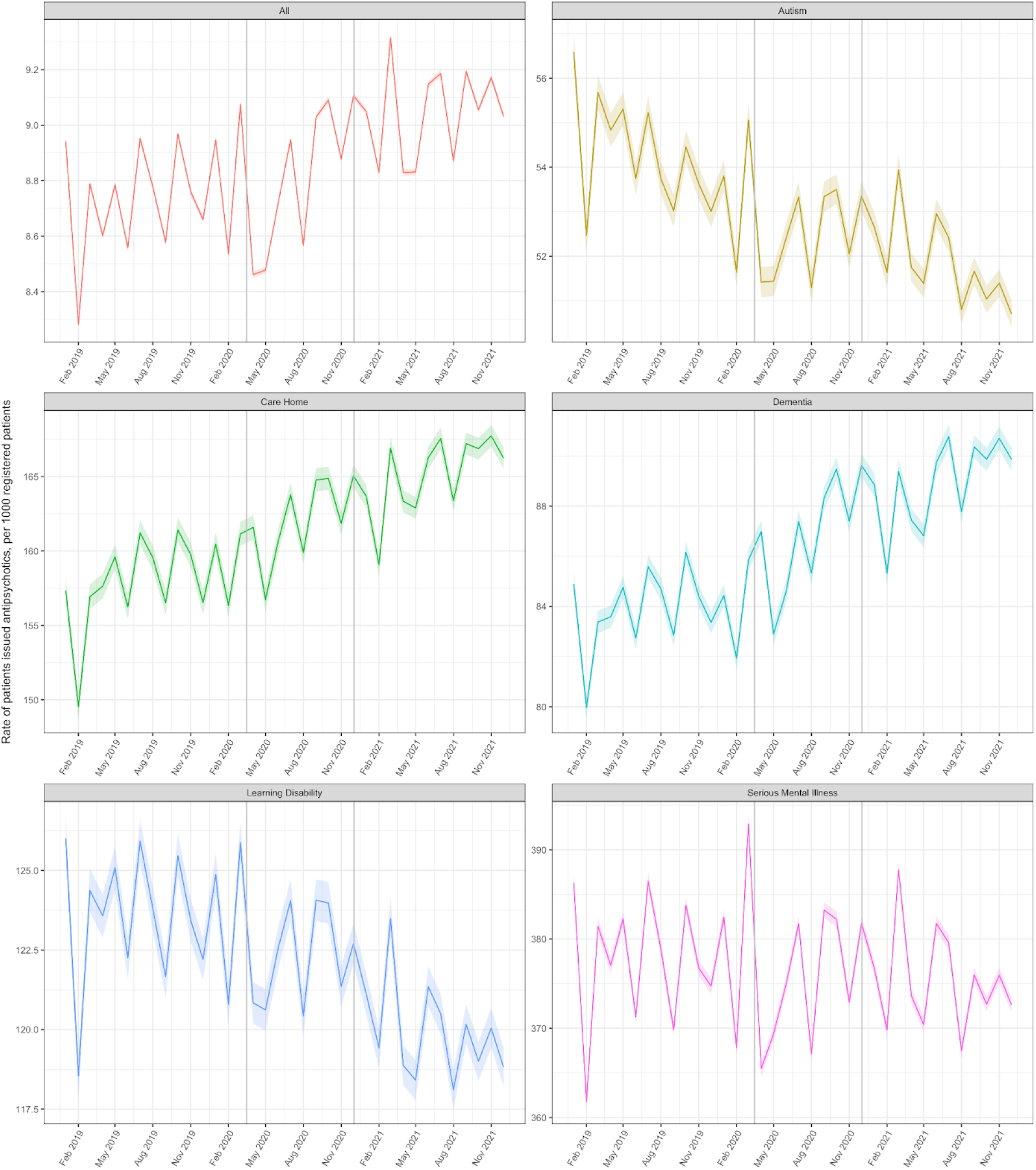
Monthly rate of patients issued an antipsychotic between January 2019 and December 2021, stratified by at-risk group. Solid coloured lines represent the monthly rates for each group with shaded colours areas representing 95% CIs. Vertical grey lines represent the start of the first two national lockdowns. Note each plot has a seperate y-axis scale thus plots should be considered separately.

**Figure 2.**
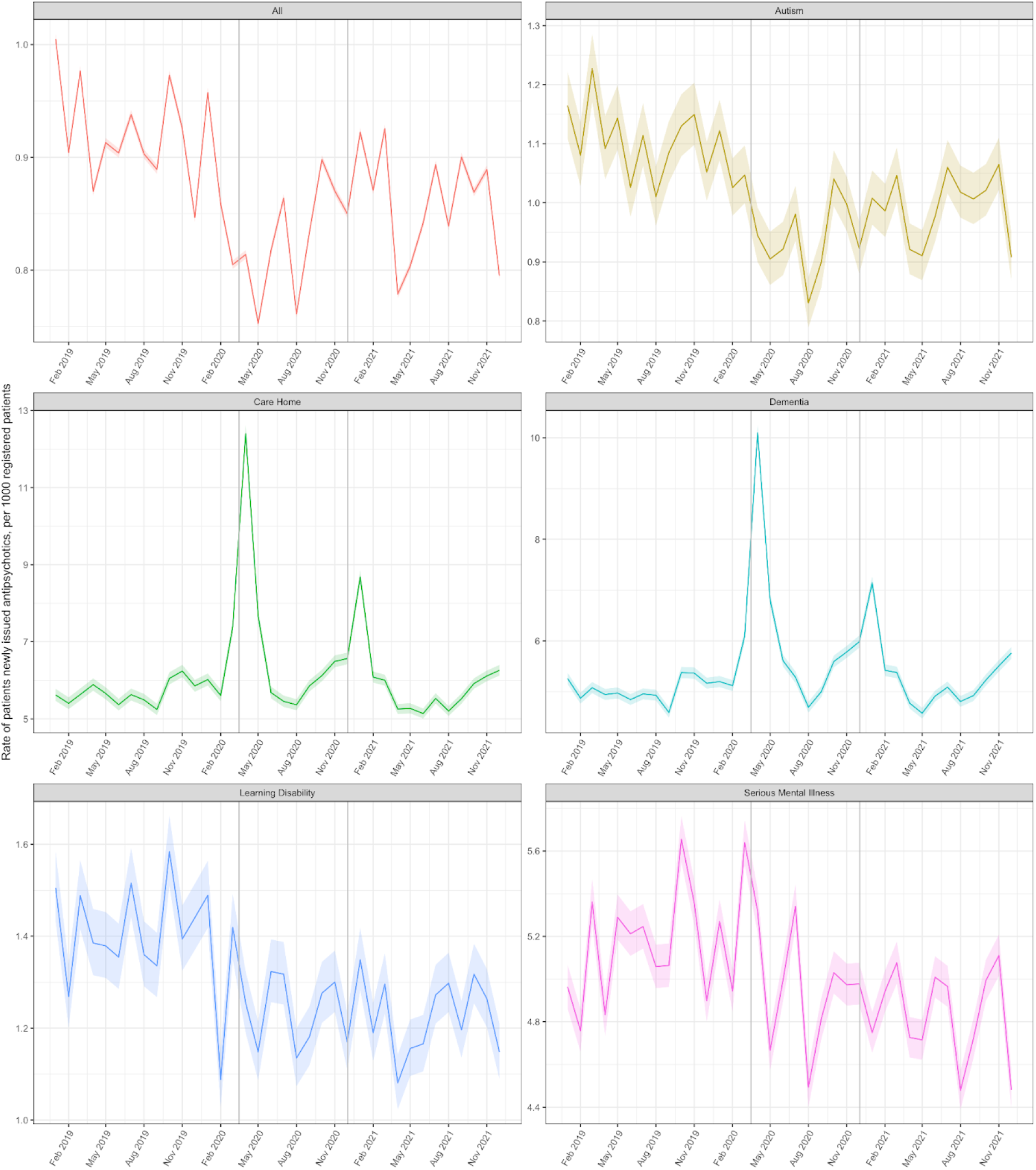
Monthly rate of patients newly issued an antipsychotic between January 2019 and December 2021 stratified by at-risk group. Solid coloured lines represent the monthly rates for each group with shaded colours areas represeting 95% CIs. Vertical grey lines represent the start of the first two national lockdowns. Note variation in each plot has a seperate y-axis scale thus means each plots should be considered accordingly.

### Demographic and Temporal variations in prescribing according to at-risk population

The populations of each group is shown in Figure 3. Further details on demographic and ethnic variabilities within the groups are shown in the Appendix in Tables S5-S9.

**Figure 3.**
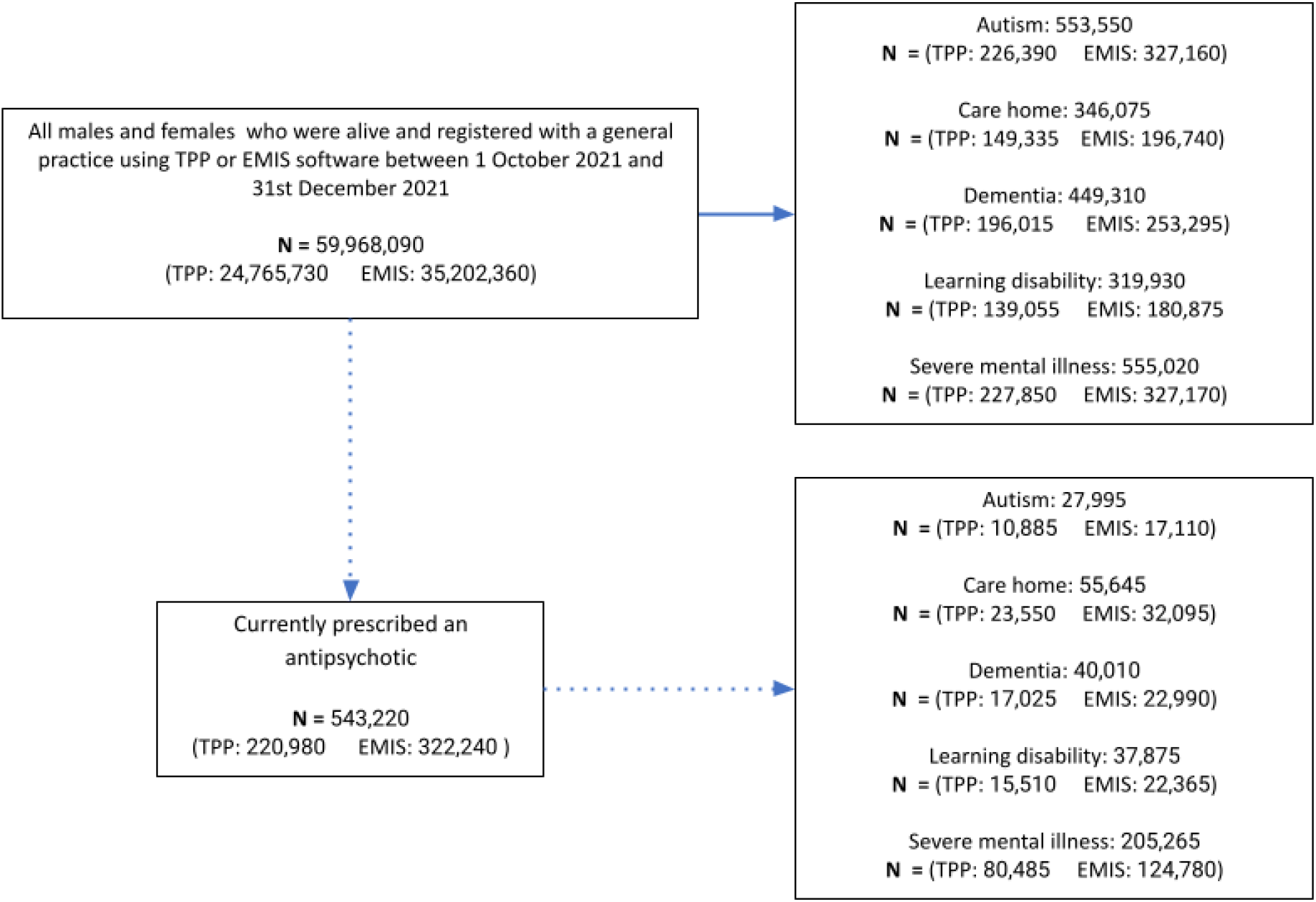
Clinical Characteristics.

### Dementia

Between 1^st^ October and 31^st^ December 2021, 449,310 individuals had a record of dementia (Figure 3). Within this population, 40,010 (9%) patients were prescribed an antipsychotic. The monthly rate (per 1000 patients) of antipsychotic prescribing increased over the study period from 82.75 (95% CI 82.30-83.19) in Q1 (Jan-Mar) 2019 to 90.1 (95% CI 89.68-90.60) in Q4 (Oct-Dec) 2021 (Figure 1).

### Care home

Between 1^st^ October and 31^st^ December 2021, 346,075 individuals were classed as residing in a care home (Figure 3). Within this population, 55,645 (16%) were prescribed an antipsychotic. The monthly rate (per 1000 patients) of antipsychotic prescribing increased over the study period from 154.61 (95% CI 153.79-155.43) in Q1 2019 to 166.95 (95% CI 166.23-167.67) in Q4 2021 (Figure 1).

### Learning disability

Between 1^st^ October and 31^st^ December 2021, 319,930 individuals were classed as having a learning disability (Figure 3). Within this population, 37,875 (12%) were prescribed an antipsychotic. The monthly rate (per 1000 patients) of antipsychotic prescribing decreased over the study period from 122.97 (95% CI 122.29-123.66) in Q1 2019 to 119.29 (95% CI 118.68-119.91) in Q4 2021 (Figure 1).

### Autism

Between 1^st^ October and 31^st^ December 2021, 553,550 individuals had a record of autism (Figure 3). From within this population, 27,995 (5%) were prescribed an antipsychotic. The monthly rate (per 1000 patients) of antipsychotic prescribing decreased slightly over the study period from 54.91 (95% CI 54.52-55.29) in Q1 2019 to 51.04 (95% CI 50.74-51.35) in Q4 2021 (Figure 1).

### Serious mental illness

Between 1^st^ October and 31^st^ December 2021, 555,020 individuals had a record of a serious mental illness (Figure 3). From within this population, 205,265 (37%) were currently prescribed an antipsychotic (Supplementary Table S5). The monthly rate (per 1000 patients) of antipsychotic prescribing within these patients decreased slightly from 376.49 (95%CI 375.60-377.38) in Q1 2019 to 373.75 (95%CI 372.92-374.57) in Q4 2021 (Figure 1).

### Prescribing trends of new prescriptions over time within at-risk populations

#### Dementia and Care Homes

Over the study period, there were two notable increases in the rate of new antipsychotic prescriptions issued to patients with dementia, which coincided with the start of the coronavirus pandemic/lockdowns; the rate of new prescriptions issued to patients with dementia rose from 5.26 (95% CI 5.14-5.37) in January 2020 to 10.09 (95% CI 9.94-10.24) in April 2020 and from 5.98 (95% CI 5.86-6.10) in November 2020 to 7.14 (95% CI 7.01-7.27) in December 2020 (Figure 2). A similar trend was seen in care homes where the rate of new prescriptions rose from 5.62 (95% CI 5.46-5.76) in January 2020 to 12.40 (95% CI 12.17-12.62) in April 2020 and from 6.56 (95% CI 6.41-6.71) in December 2020 to 8.68 (95% CI 8.51-8.50) in January 2021 (Figure 2). Our sensitivity analysis showed that when patients who were most likely to be on an palliative or end-of-life pathway were excluded, these two peaks were notably reduced (Figure 4)).

**Figure 4:**
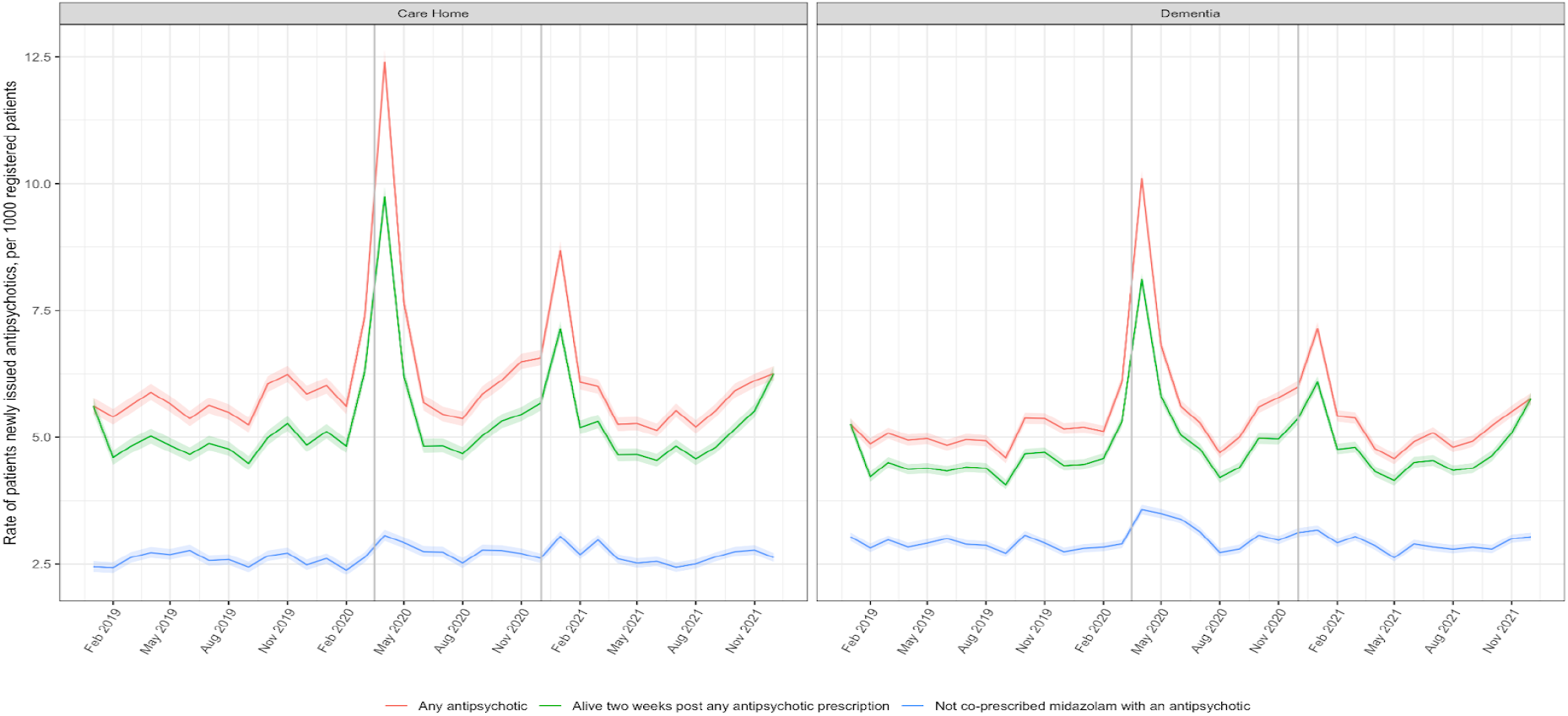
Monthly rates of care home and dementia patients **newly issued** an antipsychotic (red line), excluding patients who had died within two weeks of being prescribed an antipsychotic (one indicator of palliative care) (green line) and who had also recently been prescribed midazolam (a drug frequently prescribed in palliative care) (blue line).

#### Learning disability and Autism

The rate of initiating new antipsychotic prescriptions in patients with learning disability and in autistic patients appears to have decreased slightly over the study period, from 1.42 in Q1 2019 (95%CI 1.35-1.50) to 1.24 (95%CI 1.18-1.31) in Q4 2021 in learning disability and 1.15 (95%CI 1.10-1.21) in Q1 2019 to 0.97 (95%CI 0.96-1.04) in Q4 2021, which reflects the decrease in overall use.

#### Serious mental illness

The rate of new antipsychotic prescriptions issued to patients with serious mental illnesses remained relatively unchanged; 5.03 (95%CI 4.93-.513) in Q1 2019 and 4.86 (95%CI 4.77-4.96) in Q4 2021. Except for a slight increase in the rate of new first-generation antipsychotic prescribing in patients with serious mental illness between March and May 2020, there were no other notable trends during the course of the pandemic (Supplementary Figure S10).

## DISCUSSION

### Summary

Over the study period, we observed increases in the rate of antipsychotic prescribing in patients with dementia and in care homes. Furthermore, there were notable peaks in the rate of newly prescribed antipsychotics in these two groups which coincided with the beginning of the first and second lockdowns. Many of these new prescriptions are likely to be associated with palliative care indications. Haloperidol is recommended for the management of restlessness, confusion and vomiting during end of life care [19] and preemptive prescribing of palliative care medicines, including antipsychotics, was recommended by some organisations for elderly and frail patients in residential care, to allow staff to be ready to use it should the patient become infected with Covid [20]. Results from our sensitivity analysis are consistent with the suggestion that these spikes are due to palliative care prescribing. Conversely we observed a decreasing trend during the same period in the prescribing of antipsychotics in patients with learning disability or autism, although we found some notable variations in frequency of use associated with ethnicity which requires further investigation. We did not observe an increase in new prescriptions for antipsychotics in the learning disability and autistic population and found no consistent changes in the prescribing trends of antipsychotics in patients with a severe mental illness.

#### Strengths and weaknesses

The key strength of this study is its scale; the OpenSAFELY platform runs analyses across the full dataset of all raw, pseudonymised, single-event-level clinical events for 59.9 million patients registered at all NHS GP practices in England using EMIS and TPP software. This federated analytics allows us to explore in detail medication usage, diagnostic events, and other salient clinical, regional and demographic information including ethnicity, age and scores of deprivation. Using tailor-made codelists, we were able to design further flexibility into our data extraction which allowed for a more accurate understanding of the prescribing trends.

We also note some limitations. Whilst we have included some pre-pandemic data in our analysis, this is limited and therefore we urge caution in interpreting prescribing trends as being associated with the pandemic [21]. Some of the conditions that we were exploring are reported to not be well coded in primary care records [22]. For example in April 2021, NHS digital estimated that only 61.7% of patients with dementia had a recorded code for dementia within their GP notes [23]. It is likely that we have not included all patients in the country that have dementia and there is therefore some uncertainty about whether the identified population is fully representative of the dementia population as a whole. Furthermore, our previous work has shown that there is currently no canonical data source to identify individuals resident in a care home; and that all methods used for identifying care home residents will likely represent an under-estimate; these limitations will apply to all UK healthcare database studies [16]. All clinical groups were assessed independently, we recognise that there is significant overlap between groups such as our care home population and those with dementia or learning disability. Further analyses to explore the detail of this overlap was outside the scope of this study. We did not observe an increase in new prescriptions within the learning disability or autism cohorts, yet this must be understood within the context of our study definition: a new prescription was defined as ‘an antipsychotic prescription in those without any antipsychotic prescription in the previous 24 months’. This means that if a patient had previously taken an antipsychotic within the 2 year time period, and this antipsychotic was restarted or the dose was increased during lockdown we would not have included this in our count of new prescriptions. As a federated analysis has been carried out across two EHR vendor’s systems, it is possible that a very small number of patient records are duplicated; however, this has now been established to represent <0.03% of the total patient count. It is also possible that we counted some patients twice, if they moved GP practice and changed EHR vendor during the study period. Finally, this data only includes antipsychotics which are prescribed in primary care. In our experience some antipsychotics are prescribed directly from secondary care in some regions [24]; this includes clozapine and, in some areas, the more expensive antipsychotic injections such as Xeplion, Zypadhera, Trevicta and Abilify Maintena. Furthermore, some antipsychotics may be prescribed by secondary care services for acute behavioural disturbance, for example via psychiatric outreach teams or emergency care services, and this prescribing will not be reflected in our data.

#### Findings in Context

To our knowledge, this is the first study of its scale that used NHS GP data to describe antipsychotic prescribing trends in different at-risk patient groups, during and after the Covid pandemic.

NHS Digital publishes rates of antipsychotic prescribing in dementia over the previous 12 month period, via an interactive dashboard [23] which is updated monthly. In Nov 2020, researchers using this data observed an increase in the rate of antipsychotic prescribing in dementia in the early months of the pandemic and raised concerns that this could be in response to worsened agitation and psychosis secondary to COVID-19 restrictions [25]. In Dec 2021, NHS digital reported that 9.3% of patients with dementia had been prescribed an antipsychotic over the previous 6 weeks. This is a slightly higher figure than we observed (8.99 95%CI 8.94-9.03), although we counted prescriptions issued within a defined month, rather than over six weeks, which may account for the difference. There have also been external factors affecting their data; in Oct 2021 they removed antipsychotics commonly used in palliative care from their dataset [26] without applying this change to previous time windows, and therefore perceived reductions in their prescribing rates around this time must be viewed with caution. Throughout our data extraction we excluded antipsychotics which are used exclusively in palliative care, such as the injectable formulations of levomepromazine, however we have included those which are used for both palliative care and the management of behavioural and psychiatric symptoms of dementia, such as haloperidol.

NHS digital also publishes data about antipsychotic prescribing in learning disability. This data is calculated from a cohort of patients (56% patient coverage in 2020/21) and is reported annually [27]. In their 2020/21 report they calculated that 14.8% of patients with a learning disability were prescribed an antipsychotic; 15.2% in 2019/20. These figures are greater than ours, however they count all patients who had a prescription for an antipsychotic within a 6 month period. We have been able to provide significantly more detail in our analysis; by using a bigger dataset and a monthly extraction of data we can provide a more accurate and comprehensive description of the prescribing trends in learning disability over this period. Finally, in Canada, researchers found statistically significant increases in the use of antipsychotics, benzodiazepines, antidepressants, anticonvulsants, and opioids in nursing home residents following the onset of the COVID-19 pandemic although they noted that these increases were not as pronounced in residents who had dementia. We found no UK based research studies that described antipsychotic prescribing in care homes over the pandemic.

#### Implications for policy and research

This initial descriptive piece of work has shown how we can use the OpenSAFELY platform to facilitate near real-time research of prescribing trends in the context of an evolution of healthcare services following the pandemic. The increase in antipsychotic prescribing in dementia and care home patients despite active government initiatives to prevent their overuse is concerning and warrants further investigation and action.

A key government report recently highlighted concerns about overprescribing within the NHS, noting that it may disproportionately affect those who are more vulnerable such as the elderly and those with disabilities [28]. This report called for research on overprescribing, including a specific recommendation to prioritise research into the links between overprescribing, deprivation, ethnicity and inequalities and the impact this has on the health of the population.

#### Future research

In 2021 NHS England published their Core20Plus5 document which described their commitment to reduce health inequalities across services [29]. We have reported some information in the table and supplementary sections which indicates variations being evident in different ethnic groups in the prescribing of antipsychotics, however detailed analysis and discussion is outside the scope of this manuscript. We intend to embark on dedicated work to further explore these findings and understand the extent of variability in prescribing within these vulnerable groups. This will enable better understanding of the impact of the pandemic on these vulnerable populations and add to the evidence that supports work on reducing overprescribing and health inequalities.

Using the OpenSAFELY framework we can conduct rapid near real-time research into prescribing trends of medicines for almost the entire population of England. We can then focus in detail over a range of key variables, including medication type, diagnosis, demographics and ethnicity. With appropriate permissions and where appropriate support can be obtained from relevant professional bodies, the OpenSAFELY platform is also technically capable of providing audit and feedback information about clinical practice, and changes in clinical practice, at single sites.

#### Summary

There is still much to learn about how the pandemic and subsequent lockdowns have affected the mental health of at-risk populations. We have shown that over the course of the pandemic, there have been increases in antipsychotic use in certain at-risk populations, specifically patients with dementia and those who reside in care homes. We have also shown that there were significant brief increases in antipsychotic prescribing in patients with dementia or in those who were resident in care homes during each of the lockdown periods which are likely to be due to palliative care prescribing. Meanwhile there has been a decrease in antipsychotic prescribing in patients with learning disability or autism. We need to continue to promote and support research and policy development that reduce inappropriate prescribing of antipsychotics within these vulnerable groups of patients.

## Data Availability

All data produced in the study are contained in the manuscript and supplementary sections. All code for the OpenSAFELY platform for data management, analysis and secure code execution is shared for review and re-use under open licences on GitHub:
https://github.com/opensafely/antipsychotics-prescribing-during-COVID-19.

https://github.com/opensafely/antipsychotics-prescribing-during-COVID-19

## ADMINISTRATIVE

## Acknowledgements

We are very grateful for all the support received from the EMIS and TPP Technical Operations team throughout this work, and for generous assistance from the information governance and database teams at NHS England and the NHS England Transformation Directorate.

## Information governance and ethical approval

NHS England is the data controller; EMIS and TPP are the data processors; and the key researchers on OpenSAFELY are acting on behalf of NHS England. This implementation of OpenSAFELY is hosted within the EMIS and TPP environments which are accredited to the ISO 27001 information security standard and are NHS IG Toolkit compliant;[30,31] patient data has been pseudonymised for analysis and linkage using industry standard cryptographic hashing techniques; all pseudonymised datasets transmitted for linkage onto OpenSAFELY are encrypted; access to the platform is via a virtual private network (VPN) connection, restricted to a small group of researchers; the researchers hold contracts with NHS England and only access the platform to initiate database queries and statistical models; all database activity is logged; only aggregate statistical outputs leave the platform environment following best practice for anonymisation of results such as statistical disclosure control for low cell counts.[32] The OpenSAFELY research platform adheres to the obligations of the UK General Data Protection Regulation (GDPR) and the Data Protection Act 2018. In March 2020, the Secretary of State for Health and Social Care used powers under the UK Health Service (Control of Patient Information) Regulations 2002 (COPI) to require organisations to process confidential patient information for the purposes of protecting public health, providing healthcare services to the public and monitoring and managing the COVID-19 outbreak and incidents of exposure; this sets aside the requirement for patient consent.[33] Taken together, these provide the legal bases to link patient datasets on the OpenSAFELY platform. GP practices, from which the primary care data are obtained, are required to share relevant health information to support the public health response to the pandemic, and have been informed of the OpenSAFELY analytics platform. This study was approved by the Health Research Authority (REC reference 20/LO/0651) and by the LSHTM Ethics Board (reference 21863).

## Guarantor

Ben Goldacre is guarantor of the Opensafely project.

## Contributorship

Orla Macdonald, Amelia Green and Brian MacKenna are joint first authors.

## Authors’ contributions

|  |  |
| --- | --- |
| Conceptualization | OM, BMK, MG, HC |
| Data curation | OM, BMK, MG, AW, DE, CA, HC, PI, ID, JM, LF, SB, SD, TW, WH, BBC, CM |
| Formal Analysis | MG |
| Funding acquisition | BG |
| Investigation | MG, AW, DE |
| Methodology | OM, BMK, MG, HC, AW |
| Codelists | OM, BMK, AB, PI, RC |
| Project administration | BMK |
| Resources | BG |
| Supervision | BMK |
| Software | BG, AW, DE, CA, PI, ID, JM, MG, LF, SB, SD, TW, WH, BBC |
| Validation | AW, HC |
| Visualization | OM BMK MG |
| Writing:original draft | OM MG |
| Writing:review & edit | OM BMK MG HC AW AM |
| Information Governance | BMK AM |

