## Supplementary Material for "The impact of the COVID-19 pandemic on Antipsychotic Prescribing in individuals with autism, dementia, learning disability, serious mental illness or living in a care home: A federated analysis of 59 million patients’ primary care records in situ using OpenSAFELY"

#### Codelists

##### Table S1: Antipsychotics

| Antipsychotic medication | Codelists |
| --- | --- |
| First generation antipsychotics, excluding long acting depots | https://codelists.opensafely.org/codelist/opensafely/first-generation-antipsychotics-excluding-long-acting-depots-dmd/1e9b227c/ |
| Second generation antipsychotics excluding long acting injections | https://codelists.opensafely.org/codelist/opensafely/second-generation-antipsychotics-excluding-long-acting-injections/6c7c3c11/ |
| Long acting injectable and depot antipsychotics | https://codelists.opensafely.org/codelist/opensafely/long-acting-injectable-and-depot-antipsychotics-dmd/536cc8dc/ |
| Prochlorperazine | https://codelists.opensafely.org/codelist/opensafely/prochlorperazine-dmd/058baf47/ |

##### Table S2: At risk groups

| At risk group | Codelists |
| --- | --- |
| Learning disability | https://www.opencodelists.org/codelist/nhsd-primary-care-domain-refsets/ld_cod/20210127/ |
| Autism | https://codelists.opensafely.org/codelist/nhsd-primary-care-domain-refsets/autism_cod/20210127/ |
| Serious Mental Illness | https://www.opencodelists.org/codelist/nhsd-primary-care-domain-refsets/mh_cod/20210127/ |
| Care homes | https://www.opencodelists.org/codelist/primis-covid19-vacc-uptake/longres/v1/ |
| Dementia | https://www.opencodelists.org/codelist/nhsd-primary-care-domain-refsets/dem_cod/20210127/ |

**Table S3 Antipsychotics included in the study, according to group. Further details about products included within each codelist can be found at https://codelists.opensafely.org/**

| First Generation Antipsychotics | Second Generation Antipsychotics | Long acting injectable antipsychotics | Prochlorperazine |
|---|---|---|---|
| Benperidol<br>Chlorpromazine<br>Flupentixol<br>Haloperidol<br>Levomepromazine<br>Loxapine<br>Pericyazine<br>Pimozide<br>Promazine<br>Sulpiride<br>Trifluoperazine<br>Zuclopenthixol | Amisulpride<br>Aripiprazole<br>Asenapine<br>Cariprazine<br>Clozapine<br>Lurasidone<br>Olanzapine<br>Paliperidone<br>Quetiapine<br>Risperidone | Aripiprazole<br>Flupentixol decanoate<br>Fluphenazine decanoate<br>Haloperidol decanoate<br>Olanzapine embonate<br>Paliperidone<br>Piportil Depot<br>Risperidone<br>Zuclopenthixol decanoate | Prochlorperazine |

**Table S4 (See Table 1 in manuscript)** Characteristics of patients registered showing detail of TPP or EMIS origin. Note, this data indicates the number of patients who have received a prescription for an antipsychotic, rather than the number of prescriptions issued. Counts are rounded to the nearest 5.

| Attribute | Category | TPP | | | EMIS | | | Combined | | |
|---|---|---|---|---|---|---|---|---|---|---|
| | | Total (%) | Not on anti-psychotic (%) | Taking anti-psychotic (%) | Total (%) | Not on anti-psychotic (%) | Taking anti-psychotic (%) | Total (%) | Not on anti-psychotic (%) | Taking anti-psychotic (%)) |
| **Total** | | 24,765,730 (100) | 24,544,750 (100) | 220,980 (100) | 35,202,360 (100) | 34,880,130 (100) | 322,240 (100) | 59,968,090 (100) | 59,424,880 (100) | 543,220 (100) |
| **Age group** | **0-17** | 4,644,690 (19) | 4,642,945 (19) | 1,750 (1) | 6,638,575 (19) | 6,635,715 (19) | 2,860 (1) | 11,283,265 (19) | 11,278,660 (19) | 4,610 (1) |
| | **18-24** | 1,920,725 (8) | 1,912,180 (8) | 8,545 (4) | 2,946,405 (8) | 2,932,980 (8) | 13,425 (4) | 4,867,130 (8) | 4,845,160 (8) | 21,970 (4) |
| | **25-34** | 3,421,385 (14) | 3,395,605 (14) | 25,780 (12) | 5,225,690 (15) | 5,188,745 (15) | 36,950 (11) | 8,647,075 (14) | 8,584,350 (14) | 62,730 (12) |
| | **35-44** | 3,440,625 (14) | 3,407,630 (14) | 32,995 (15) | 5,102,660 (14) | 5,054,590 (14) | 48,070 (15) | 8,543,285 (14) | 8,462,220 (14) | 81,065 (15) |
| | **45-54** | 3,214,825 (13) | 3,175,245 (13) | 39,580 (18) | 4,552,070 (13) | 4,492,890 (13) | 59,185 (18) | 7,766,895 (13) | 7,668,135 (13) | 98,765 (18) |
| | **55-69** | 4,473,325 (18) | 4,417,190 (18) | 56,135 (25) | 6,032,040 (17) | 5,947,765 (17) | 84,275 (26) | 10,505,365 (18) | 10,364,955 (17) | 140,410 (26) |
| | **70-79** | 2,236,870 (9) | 2,208,410 (9) | 28,460 (13) | 2,862,400 (8) | 2,822,735 (8) | 39,665 (12) | 5,099,270 (9) | 5,031,145 (8) | 68,125 (13) |
| | **80+** | 1,413,285 (6) | 1,385,545 (6) | 27,735 (13) | 1,842,520 (5) | 1,804,710 (5) | 37,810 (12) | 3,255,805 (5) | 3,190,255 (5) | 65,545 (12) |
| **Sex** | **Female** | 12,367,160 (50) | 12,239,700 (50) | 127,460 (58) | 17,570,810 (50) | 17,387,880 (50) | 182,930 (57) | 29,937,970 (50) | 29,627,580 (50) | 310,390 (57) |
| | **Male** | 12,398,575 (50) | 12,305,050 (50) | 93,525 (42) | 17,631,555 (50) | 17,492,250 (50) | 139,300 (43) | 30,030,130 (50) | 29,797,300 (50) | 232,825 (43) |
| **IMD** | **1 Most deprived** | 4,923,985 (20) | 4,862,120 (20) | 61,860 (28) | 7,254,655 (21) | 7,156,750 (21) | 97,905 (30) | 12,178,640 (20) | 12,018,870 (20) | 159,765 (29) |
| | **2** | 4,835,865 (20) | 4,785,965 (19) | 49,895 (23) | 7,540,985 (21) | 7,465,530 (21) | 75,455 (23) | 12,376,850 (21) | 12,251,495 (21) | 125,350 (23) |
| | **3** | 5,087,635 (21) | 5,043,835 (21) | 43,805 (20) | 6,850,980 (19) | 6,793,105 (19) | 57,875 (18) | 11,938,615 (20) | 11,836,940 (20) | 101,680 (19) |
| | **4** | 4,784,120 (19) | 4,750,065 (19) | 34,055 (15) | 6,587,990 (19) | 6,540,025 (19) | 47,965 (15) | 11,372,110 (19) | 11,290,090 (19) | 82,020 (15) |
| | **5 Least deprived** | 4,375,810 (18) | 4,350,135 (18) | 25,675 (12) | 6,841,460 (19) | 6,800,240 (19) | 41,220 (13) | 11,217,270 (19) | 11,150,375 (19) | 66,895 (12) |
| | **Unknown** | 758,320 (3) | 752,635 (3) | 5,690 (3) | 126,295 (0) | 124,480 (0) | 1,815 (1) | 884,615 (1) | 877,115 (1) | 7,505 (1) |
| **Ethnicity** | **African** | 285,070 (1) | 283,305 (1) | 1,770 (1) | 776,310 (2) | 770,575 (2) | 5,735 (2) | 1,061,380 (2) | 1,053,880 (2) | 7,505 (1) |
| | **Bangladeshi or British Bangladeshi** | 99,360 (0) | 98,510 (0) | 850 (0) | 375,240 (1) | 371,505 (1) | 3,735 (1) | 474,600 (1) | 470,015 (1) | 4,585 (1) |
| | **British or Mixed British** | 10,311,070 (42) | 10,190,620 (42) | 120,445 (55) | 16,233,665 (46) | 16,038,615 (46) | 195,050 (61) | 26,544,735 (44) | 26,229,235 (44) | 315,495 (58) |
| | **Caribbean** | 84,410 (0) | 83,120 (0) | 1,290 (1) | 291,200 (1) | 286,920 (1) | 4,280 (1) | 375,610 (1) | 370,040 (1) | 5,570 (1) |
| | **Chinese** | 140,635 (1) | 140,290 (1) | 340 (0) | 344,750 (1) | 344,030 (1) | 720 (0) | 485,385 (1) | 484,320 (1) | 1,060 (0) |
| | **Indian or British Indian** | 566,770 (2) | 563,250 (2) | 3,515 (2) | 981,195 (3) | 975,245 (3) | 5,945 (2) | 1,547,965 (3) | 1,538,495 (3) | 9,460 (2) |
| | **Irish** | 83,845 (0) | 82,635 (0) | 1,210 (1) | 192,815 (1) | 189,860 (1) | 2,960 (1) | 276,660 (0) | 272,495 (0) | 4,170 (1) |

| | | | | | | | | | | |
|---|---|---|---|---|---|---|---|---|---|---|
| | **Other** | 294,655 (1) | 292,655 (1) | 2,000 (1) | 551,010 (2) | 547,465 (2) | 3,545 (1) | 845,665 (1) | 840,120 (1) | 5,545 (1) |
| | **Other Asian** | 308,105 (1) | 306,275 (1) | 1,825 (1) | 667,035 (2) | 662,685 (2) | 4,350 (1) | 975,140 (2) | 968,960 (2) | 6,175 (1) |
| | **Other Black** | 68,855 (0) | 68,215 (0) | 640 (0) | 201,755 (1) | 199,495 (1) | 2,255 (1) | 270,610 (0) | 267,710 (0) | 2,895 (1) |
| | **Other mixed** | 108,565 (0) | 107,830 (0) | 735 (0) | 266,700 (1) | 264,935 (1) | 1,765 (1) | 375,265 (1) | 372,765 (1) | 2,500 (0) |
| | **Other White** | 1,512,220 (6) | 1,506,095 (6) | 6,125 (3) | 2,695,825 (8) | 2,682,935 (8) | 12,885 (4) | 4,208,045 (7) | 4,189,030 (7) | 19,010 (3) |
| | **Pakistani or British Pakistani** | 434,565 (2) | 430,580 (2) | 3,990 (2) | 772,290 (2) | 765,565 (2) | 6,725 (2) | 1,206,855 (2) | 1,196,145 (2) | 10,715 (2) |
| | **White + Asian** | 68,575 (0) | 68,165 (0) | 410 (0) | 124,130 (0) | 123,350 (0) | 780 (0) | 192,705 (0) | 191,515 (0) | 1,190 (0) |
| | **White + Black African** | 66,545 (0) | 66,105 (0) | 440 (0) | 130,430 (0) | 129,580 (0) | 850 (0) | 196,975 (0) | 195,685 (0) | 1,290 (0) |
| | **White + Black Caribbean** | 70,975 (0) | 70,125 (0) | 850 (0) | 142,720 (0) | 140,985 (0) | 1,735 (1) | 213,695 (0) | 211,110 (0) | 2,585 (0) |
| | **Unknown** | 10,261,515 (41) | 10,186,970 (42) | 74,540 (34) | 10,455,295 (30) | 10,386,380 (30) | 68,915 (21) | 20,716,810 (35) | 20,573,350 (35) | 143,455 (26) |
| **Region** | **East of England** | 5,767,745 (23) | 5,711,595 (23) | 56,150 (25) | 1,308,015 (4) | 1,296,415 (4) | 11,600 (4) | 7,075,760 (12) | 7,008,010 (12) | 67,750 (12) |
| | **London** | 1,812,115 (7) | 1,800,535 (7) | 11,580 (5) | 8,310,930 (24) | 8,247,590 (24) | 63,340 (20) | 10,123,045 (17) | 10,048,125 (17) | 74,920 (14) |
| | **Midlands** | 5,277,845 (21) | 5,227,650 (21) | 50,195 (23) | 5,990,605 (17) | 5,930,230 (17) | 60,370 (19) | 11,268,450 (19) | 11,157,880 (19) | 110,565 (20) |
| | **North East and Yorkshire** | 4,707,565 (19) | 4,666,430 (19) | 41,135 (19) | 2,733,920 (8) | 2,709,805 (8) | 24,115 (7) | 7,441,485 (12) | 7,376,235 (12) | 65,250 (12) |
| | **North West** | 2,115,735 (9) | 2,098,975 (9) | 16,760 (8) | 6,844,105 (19) | 6,768,540 (19) | 75,565 (23) | 8,959,840 (15) | 8,867,515 (15) | 92,325 (17) |
| | **South East** | 1,656,560 (7) | 1,641,305 (7) | 15,255 (7) | 7,452,010 (21) | 7,386,880 (21) | 65,130 (20) | 9,108,570 (15) | 9,028,185 (15) | 80,385 (15) |
| | **South West** | 3,393,385 (14) | 3,363,830 (14) | 29,560 (13) | 2,562,780 (7) | 2,540,665 (7) | 22,110 (7) | 5,956,165 (10) | 5,904,495 (10) | 51,670 (10) |
| | **Unknown** | 34,785 (0) | 34,445 (0) | 345 (0) | 0 (0) | 0 (0) | 0 (0) | 34,785 (0) | 34,445 (0) | 345 (0) |

**Table S5:** Counts and rates of dementia patients currently prescribed an antipsychotic between 1st October 2021 and 31st December 2021, stratified by demographic variables

| Attribute | Category | TPP | | EMIS | | Combined | | | |
|---|---|---|---|---|---|---|---|---|---|
| | | Total (%) | Rate per 1000 | Total (%) | Rate per 1000 | Total (%) | Rate per 1000 | Lower 95% CI | Upper 95% CI |
| Total | | 17,030 (9) | 86.88 | 22,990 (9) | 90.76 | 40,020 (9) | 89.07 | 88.20 | 89.94 |
| Age group | 0-17 | - | - | - | - | - | - | - | - |
| | 18-24 | - | - | - | - | - | - | - | - |
| | 25-34 | - | - | - | - | - | - | - | - |
| | 35-44 | 10 (11) | 111.11 | 15 (23) | 230.77 | 25 (16) | 161.29 | 98.07 | 224.51 |
| | 45-54 | 105 (15) | 147.89 | 160 (20) | 202.53 | 265 (18) | 176.67 | 155.40 | 197.94 |
| | 55-69 | 1,525 (14) | 144.28 | 2,245 (17) | 165.44 | 3,770 (16) | 156.17 | 151.19 | 161.16 |
| | 70-79 | 4,690 (11) | 113.79 | 6,370 (12) | 122.19 | 11,060 (12) | 118.49 | 116.28 | 120.69 |
| | 80+ | 10,700 (7) | 74.60 | 14,195 (8) | 76.02 | 24,895 (8) | 75.40 | 74.47 | 76.34 |
| Sex | Female | 10,540 (9) | 85.86 | 14,310 (9) | 90.04 | 24,850 (9) | 88.22 | 87.12 | 89.31 |
| | Male | 6,485 (9) | 88.53 | 8,675 (9) | 91.93 | 15,160 (9) | 90.44 | 89.00 | 91.88 |
| IMD | 1 Most deprived | 3,465 (10) | 101.97 | 4,635 (10) | 103.33 | 8,100 (10) | 102.75 | 100.51 | 104.98 |
| | 2 | 3,485 (9) | 94.06 | 4,540 (10) | 95.24 | 8,025 (9) | 94.72 | 92.65 | 96.80 |
| | 3 | 3,800 (9) | 88.28 | 4,720 (9) | 93.29 | 8,520 (9) | 90.99 | 89.05 | 92.92 |
| | 4 | 3,115 (8) | 75.99 | 4,685 (9) | 86.85 | 7,800 (8) | 82.16 | 80.34 | 83.98 |
| | 5 Least deprived | 2,800 (7) | 74.96 | 4,345 (8) | 78.22 | 7,145 (8) | 76.91 | 75.12 | 78.69 |
| | Unknown | 360 (10) | 100.14 | 65 (10) | 95.59 | 425 (10) | 99.42 | 89.96 | 108.87 |
| Ethnicity | African | 35 (8) | 75.27 | 145 (9) | 88.69 | 180 (9) | 85.71 | 73.19 | 98.24 |
| | Bangladeshi or British Bangladeshi | 25 (9) | 89.29 | 100 (10) | 98.52 | 125 (10) | 96.53 | 79.60 | 113.45 |
| | British or Mixed British | 9,320 (9) | 90.33 | 15,095 (9) | 94.56 | 24,415 (9) | 92.90 | 91.73 | 94.06 |
| | Caribbean | 100 (9) | 86.21 | 335 (8) | 83.44 | 435 (8) | 84.06 | 76.16 | 91.96 |
| | Chinese | 15 (9) | 85.71 | 30 (7) | 70.59 | 45 (8) | 75.00 | 53.09 | 96.91 |
| | Indian or British Indian | 150 (7) | 70.75 | 345 (9) | 92.74 | 495 (8) | 84.76 | 77.29 | 92.23 |
| | Irish | 145 (10) | 100.35 | 335 (11) | 105.85 | 480 (10) | 104.12 | 94.81 | 113.44 |
| | Other | 55 (9) | 90.91 | 130 (12) | 119.27 | 185 (11) | 109.14 | 93.42 | 124.87 |
| | Other Asian | 50 (7) | 72.46 | 135 (8) | 78.72 | 185 (8) | 76.92 | 65.84 | 88.01 |
| | Other Black | 15 (9) | 90.91 | 55 (10) | 101.85 | 70 (10) | 99.29 | 76.03 | 122.55 |
| | Other mixed | 15 (9) | 93.75 | 40 (10) | 95.24 | 55 (9) | 94.83 | 69.77 | 119.89 |

| | | | | | | | | | |
|---|---|---|---|---|---|---|---|---|---|
| | Other White | 285 (8) | 84.32 | 610 (9) | 87.77 | 895 (9) | 86.64 | 80.96 | 92.32 |
| | Pakistani or British Pakistani | 125 (9) | 92.94 | 200 (9) | 94.34 | 325 (9) | 93.80 | 83.60 | 103.99 |
| | Unknown | 6,640 (8) | 82.62 | 5,335 (8) | 80.97 | 11,975 (8) | 81.88 | 80.41 | 83.35 |
| | White + Asian | 10 (9) | 90.91 | 20 (11) | 114.29 | 30 (11) | 105.26 | 67.60 | 142.93 |
| | White + Black African | 15 (18) | 176.47 | 20 (10) | 97.56 | 35 (12) | 120.69 | 80.71 | 160.67 |
| | White + Black Caribbean | 25 (8) | 84.75 | 60 (10) | 102.56 | 85 (10) | 96.59 | 76.06 | 117.12 |
| **Region** | East of England | 4,500 (10) | 102.03 | 1,165 (11) | 113.38 | 5,665 (10) | 104.17 | 101.46 | 106.89 |
| | London | 445 (6) | 61.21 | 3,495 (8) | 84.86 | 3,940 (8) | 81.31 | 78.77 | 83.85 |
| | Midlands | 3,940 (9) | 93.89 | 4,720 (10) | 104.41 | 8,660 (10) | 99.35 | 97.25 | 101.44 |
| | North East and Yorkshire | 3,305 (9) | 88.10 | 1,630 (8) | 76.45 | 4,935 (8) | 83.88 | 81.54 | 86.22 |
| | North West | 1,220 (6) | 64.35 | 4,585 (8) | 84.93 | 5,805 (8) | 79.58 | 77.53 | 81.63 |
| | South East | 1,165 (8) | 78.14 | 5,525 (9) | 92.66 | 6,690 (9) | 89.76 | 87.61 | 91.91 |
| | South West | 2,430 (8) | 78.10 | 1,865 (9) | 85.94 | 4,295 (8) | 81.32 | 78.89 | 83.75 |
| | Unknown | 20 (11) | 111.11 | 0 (0) | 0 (0) | 20 (11) | 111.11 | 62.42 | 159.81 |

**Table S6:** Counts and rates of care home patients currently prescribed an antipsychotic between 1st October 2021 and 31st December 2021, stratified by demographic variables

| Attribute | Category | TPP | | EMIS | | Combined | | | |
|---|---|---|---|---|---|---|---|---|---|
| | | Total (%) | Rate per 1000 | Total (%) | Rate per 1000 | Total (%) | Rate per 1000 | Lower 95% CI | Upper 95% CI |
| Total | | 23,550 (16) | 157.70 | 32,080 (16) | 163.06 | 55,630 (16) | 160.75 | 159.41 | 162.08 |
| Age group | 0-17 | 30 (7) | 74.07 | 30 (5) | 45.45 | 60 (6) | 56.34 | 42.08 | 70.59 |
| | 18-24 | 350 (19) | 191.78 | 375 (19) | 194.30 | 725 (19) | 193.08 | 179.02 | 207.13 |
| | 25-34 | 1,150 (23) | 231.85 | 1,310 (25) | 251.20 | 2,460 (24) | 241.77 | 232.22 | 251.32 |
| | 35-44 | 1,355 (26) | 259.33 | 1,715 (30) | 296.46 | 3,070 (28) | 278.84 | 268.97 | 288.70 |
| | 45-54 | 1,945 (29) | 285.82 | 2,525 (31) | 314.64 | 4,470 (30) | 301.42 | 292.58 | 310.25 |
| | 55-69 | 4,825 (29) | 286.10 | 6,780 (31) | 308.60 | 11,605 (30) | 298.83 | 293.39 | 304.27 |
| | 70-79 | 4,760 (22) | 216.27 | 6,945 (23) | 234.75 | 11,705 (23) | 226.86 | 222.75 | 230.97 |
| | 80+ | 9,135 (10) | 100.12 | 12,405 (10) | 100.39 | 21,540 (10) | 100.27 | 98.94 | 101.61 |
| Sex | Female | 12,815 (14) | 135.48 | 17,100 (14) | 138.05 | 29,915 (14) | 136.94 | 135.38 | 138.49 |
| | Male | 10,735 (20) | 196.09 | 14,995 (21) | 205.78 | 25,730 (20) | 201.62 | 199.16 | 204.09 |
| IMD | 1 Most deprived | 5,340 (18) | 180.71 | 7,205 (19) | 185.05 | 12,545 (18) | 183.18 | 179.97 | 186.38 |
| | 2 | 5,350 (17) | 169.55 | 7,050 (18) | 178.03 | 12,400 (17) | 174.27 | 171.20 | 177.33 |
| | 3 | 5,325 (16) | 161.85 | 6,795 (17) | 169.49 | 12,120 (17) | 166.05 | 163.09 | 169.01 |
| | 4 | 4,055 (14) | 140.14 | 5,920 (15) | 145.60 | 9,975 (14) | 143.33 | 140.52 | 146.14 |
| | 5 Least deprived | 3,030 (13) | 127.77 | 5,000 (14) | 135.83 | 8,030 (13) | 132.67 | 129.77 | 135.57 |
| | Unknown | 450 (17) | 167.91 | 125 (19) | 193.80 | 575 (17) | 172.93 | 158.80 | 187.07 |
| Ethnicity | African | 70 (18) | 181.82 | 280 (24) | 238.30 | 350 (22) | 224.36 | 200.85 | 247.86 |
| | Bangladeshi or British Bangladeshi | 15 (16) | 157.89 | 100 (24) | 243.90 | 115 (23) | 227.72 | 186.10 | 269.34 |
| | British or Mixed British | 14,325 (17) | 168.91 | 22,125 (17) | 173.24 | 36,450 (17) | 171.51 | 169.75 | 173.27 |
| | Caribbean | 150 (25) | 254.24 | 525 (27) | 272.73 | 675 (27) | 268.39 | 248.14 | 288.64 |
| | Chinese | 20 (17) | 166.67 | 45 (16) | 160.71 | 65 (16) | 162.50 | 123.00 | 202.00 |
| | Indian or British Indian | 175 (21) | 207.10 | 340 (23) | 228.19 | 515 (22) | 220.56 | 201.51 | 239.61 |
| | Irish | 135 (16) | 157.89 | 345 (18) | 178.76 | 480 (17) | 172.35 | 156.93 | 187.77 |
| | Other | 85 (19) | 188.89 | 180 (20) | 204.55 | 265 (20) | 199.25 | 175.26 | 223.24 |
| | Other Asian | 90 (21) | 211.76 | 200 (21) | 208.33 | 290 (21) | 209.39 | 185.29 | 233.49 |
| | Other Black | 45 (23) | 230.77 | 220 (31) | 307.69 | 265 (29) | 291.21 | 256.15 | 326.27 |
| | Other mixed | 45 (26) | 257.14 | 80 (21) | 205.13 | 125 (22) | 221.24 | 182.45 | 260.02 |
| | Other White | 365 (14) | 140.38 | 730 (16) | 163.31 | 1,095 (15) | 154.88 | 145.71 | 164.05 |

| | | | | | | | | | |
|---|---|---|---|---|---|---|---|---|---|
| | Pakistani or British Pakistani | 115 (27) | 270.59 | 140 (26) | 261.68 | 255 (27) | 265.62 | 233.02 | 298.23 |
| | Unknown | 7,805 (14) | 137.17 | 6,580 (12) | 124.00 | 14,385 (13) | 130.81 | 128.68 | 132.95 |
| | White + Asian | 20 (18) | 181.82 | 35 (21) | 205.88 | 55 (20) | 196.43 | 144.52 | 248.34 |
| | White + Black African | 25 (23) | 227.27 | 40 (22) | 222.22 | 65 (22) | 224.14 | 169.65 | 278.63 |
| | White + Black Caribbean | 65 (28) | 276.60 | 115 (25) | 252.75 | 180 (26) | 260.87 | 222.76 | 298.98 |
| **Region** | East of England | 5,550 (17) | 170.27 | 1,485 (17) | 171.38 | 7,035 (17) | 170.50 | 166.52 | 174.49 |
| | London | 555 (16) | 155.46 | 5,150 (19) | 191.56 | 5,705 (19) | 187.33 | 182.46 | 192.19 |
| | Midlands | 5,730 (18) | 176.06 | 6,215 (18) | 177.07 | 11,945 (18) | 176.58 | 173.42 | 179.75 |
| | North East and Yorkshire | 4,705 (16) | 161.66 | 2,625 (16) | 155.60 | 7,330 (16) | 159.43 | 155.78 | 163.08 |
| | North West | 1,830 (12) | 120.04 | 6,640 (15) | 150.40 | 8,470 (14) | 142.60 | 139.57 | 145.64 |
| | South East | 1,755 (15) | 145.95 | 7,275 (15) | 154.02 | 9,030 (15) | 152.38 | 149.24 | 155.52 |
| | South West | 3,385 (14) | 140.60 | 2,705 (15) | 151.67 | 6,090 (15) | 145.31 | 141.66 | 148.96 |
| | Unknown | 35 (20) | 200.00 | 0 (0) | 0 (0) | 35 (20) | 200.00 | 133.74 | 266.26 |

**Table S7:** Counts and rates of patients with a learning disability currently prescribed an antipsychotic between 1st October 2021 and 31st December 2021, stratified by demographic variables

| Attribute | Category | TPP | | EMIS | | Combined | | | |
|---|---|---|---|---|---|---|---|---|---|
| | | Total (%) | Rate per 1000 | Total (%) | Rate per 1000 | Total (%) | Rate per 1000 | Lower 95% CI | Upper 95% CI |
| Total | | 15,505 (11) | 111.50 | 22,365 (12) | 123.65 | 37,870 (12) | 118.37 | 117.18 | 119.56 |
| Age group | 0-17 | 220 (1) | 11.66 | 330 (1) | 12.46 | 550 (1) | 12.13 | 11.11 | 13.14 |
| | 18-24 | 1,225 (6) | 59.52 | 1,820 (7) | 68.67 | 3,045 (6) | 64.67 | 62.37 | 66.97 |
| | 25-34 | 2,945 (10) | 95.40 | 4,230 (11) | 108.66 | 7,175 (10) | 102.79 | 100.42 | 105.17 |
| | 35-44 | 2,635 (13) | 125.87 | 3,665 (14) | 138.35 | 6,300 (13) | 132.84 | 129.56 | 136.12 |
| | 45-54 | 2,860 (16) | 162.36 | 3,975 (18) | 178.53 | 6,835 (17) | 171.39 | 167.33 | 175.45 |
| | 55-69 | 4,305 (19) | 188.16 | 6,255 (21) | 207.91 | 10,560 (20) | 199.38 | 195.57 | 203.18 |
| | 70-79 | 1,110 (19) | 190.72 | 1,720 (22) | 215.13 | 2,830 (20) | 204.85 | 197.30 | 212.40 |
| | 80+ | 205 (14) | 137.58 | 375 (18) | 177.73 | 580 (16) | 161.11 | 148.00 | 174.22 |
| Sex | Female | 5,860 (11) | 107.15 | 8,340 (12) | 118.74 | 14,200 (11) | 113.67 | 111.80 | 115.54 |
| | Male | 9,650 (11) | 114.38 | 14,025 (13) | 126.76 | 23,675 (12) | 121.41 | 119.86 | 122.95 |
| IMD | 1 Most deprived | 4,265 (10) | 104.46 | 6,355 (12) | 118.02 | 10,620 (11) | 112.17 | 110.04 | 114.31 |
| | 2 | 3,820 (12) | 120.41 | 5,480 (13) | 127.25 | 9,300 (12) | 124.35 | 121.82 | 126.88 |
| | 3 | 3,280 (12) | 120.17 | 4,450 (13) | 131.81 | 7,730 (13) | 126.61 | 123.78 | 129.43 |
| | 4 | 2,400 (11) | 114.45 | 3,470 (13) | 126.60 | 5,870 (12) | 121.33 | 118.23 | 124.43 |
| | 5 Least deprived | 1,435 (9) | 94.91 | 2,530 (11) | 114.69 | 3,965 (11) | 106.64 | 103.32 | 109.96 |
| | Unknown | 310 (10) | 99.52 | 80 (11) | 108.84 | 390 (10) | 101.30 | 91.25 | 111.35 |
| Ethnicity | African | 80 (8) | 82.90 | 355 (10) | 95.82 | 435 (9) | 93.15 | 84.39 | 101.90 |
| | Bangladeshi or British Bangladeshi | 60 (10) | 102.56 | 250 (11) | 105.71 | 310 (11) | 105.08 | 93.39 | 116.78 |
| | British or Mixed British | 9,840 (13) | 127.49 | 15,325 (14) | 139.14 | 25,165 (13) | 134.34 | 132.68 | 136.00 |
| | Caribbean | 105 (16) | 155.56 | 400 (17) | 169.85 | 505 (17) | 166.67 | 152.13 | 181.20 |
| | Chinese | 20 (12) | 121.21 | 35 (9) | 92.11 | 55 (10) | 100.92 | 74.25 | 127.59 |
| | Indian or British Indian | 225 (12) | 116.58 | 385 (12) | 121.45 | 610 (12) | 119.61 | 110.12 | 129.10 |
| | Irish | 50 (17) | 166.67 | 150 (18) | 182.93 | 200 (18) | 178.57 | 153.82 | 203.32 |
| | Other | 70 (10) | 95.89 | 120 (8) | 83.62 | 190 (9) | 87.76 | 75.28 | 100.24 |
| | Other Asian | 90 (9) | 92.31 | 235 (11) | 105.38 | 325 (10) | 101.40 | 90.38 | 112.43 |
| | Other Black | 55 (14) | 135.80 | 255 (14) | 138.96 | 310 (14) | 138.39 | 122.99 | 153.80 |
| | Other mixed | 45 (8) | 82.57 | 110 (9) | 90.53 | 155 (9) | 88.07 | 74.20 | 101.93 |

| | | | | | | | | | |
|---|---|---|---|---|---|---|---|---|---|
| | Other White | 240 (9) | 91.95 | 600 (11) | 114.72 | 840 (11) | 107.14 | 99.90 | 114.39 |
| | Pakistani or British Pakistani | 315 (9) | 87.02 | 485 (9) | 86.45 | 800 (9) | 86.67 | 80.67 | 92.68 |
| | Unknown | 4,185 (9) | 88.45 | 3,405 (9) | 89.11 | 7,590 (9) | 88.75 | 86.75 | 90.74 |
| | White + Asian | 35 (12) | 122.81 | 55 (11) | 105.77 | 90 (11) | 111.80 | 88.70 | 134.90 |
| | White + Black African | 20 (8) | 81.63 | 55 (10) | 95.65 | 75 (9) | 91.46 | 70.76 | 112.16 |
| | White + Black Caribbean | 70 (13) | 133.33 | 145 (13) | 133.64 | 215 (13) | 133.54 | 115.69 | 151.39 |
| **Region** | East of England | 3,555 (12) | 121.79 | 825 (13) | 129.41 | 4,380 (12) | 123.15 | 119.51 | 126.80 |
| | London | 530 (10) | 95.58 | 4,225 (12) | 118.10 | 4,755 (12) | 115.08 | 111.81 | 118.35 |
| | Midlands | 3,785 (12) | 124.42 | 4,320 (13) | 134.98 | 8,105 (13) | 129.84 | 127.01 | 132.66 |
| | North East and Yorkshire | 3,260 (11) | 107.22 | 2,130 (13) | 125.78 | 5,390 (11) | 113.86 | 110.82 | 116.90 |
| | North West | 1,175 (8) | 82.57 | 4,750 (12) | 120.13 | 5,925 (11) | 110.19 | 107.39 | 113.00 |
| | South East | 995 (11) | 105.46 | 4,545 (13) | 125.12 | 5,540 (12) | 121.07 | 117.88 | 124.25 |
| | South West | 2,190 (11) | 111.39 | 1,570 (11) | 112.79 | 3,760 (11) | 111.97 | 108.39 | 115.55 |
| | Unknown | 25 (14) | 142.86 | 0 (0) | 0 (0) | 25 (14) | 142.86 | 86.86 | 198.86 |

**Table S8:** Counts and rates of patients with autism currently prescribed an antipsychotic between 1st October 2021 and 31st December 2021, stratified by demographic variables

| Attribute | Category | TPP | | EMIS | | Combined | | | |
|---|---|---|---|---|---|---|---|---|---|
| | | Total (%) | Rate per 1000 | Total (%) | Rate per 1000 | Total (%) | Rate per 1000 | Lower 95% CI | Upper 95% CI |
| Total | | 10,885 (5) | 48.08 | 17,110 (5) | 52.30 | 27,995 (5) | 50.57 | 49.98 | 51.17 |
| Age group | 0-17 | 585 (1) | 5.57 | 1,055 (1) | 6.84 | 1,640 (1) | 6.32 | 6.02 | 6.63 |
| | 18-24 | 2,065 (4) | 39.77 | 3,455 (5) | 45.75 | 5,520 (4) | 43.32 | 42.17 | 44.46 |
| | 25-34 | 3,275 (8) | 80.22 | 5,035 (9) | 89.50 | 8,310 (9) | 85.60 | 83.75 | 87.44 |
| | 35-44 | 1,910 (14) | 142.80 | 2,825 (15) | 151.68 | 4,735 (15) | 147.97 | 143.75 | 152.18 |
| | 45-54 | 1,445 (18) | 183.26 | 2,185 (20) | 195.70 | 3,630 (19) | 190.55 | 184.35 | 196.75 |
| | 55-69 | 1,400 (22) | 218.24 | 2,175 (22) | 224.46 | 3,575 (22) | 221.98 | 214.70 | 229.26 |
| | 70-79 | 180 (22) | 223.60 | 330 (25) | 248.12 | 510 (24) | 238.88 | 218.14 | 259.61 |
| | 80+ | 25 (17) | 172.41 | 55 (22) | 220.00 | 80 (20) | 202.53 | 158.15 | 246.91 |
| Sex | Female | 3,225 (6) | 55.63 | 5,130 (6) | 60.20 | 8,355 (6) | 58.35 | 57.10 | 59.60 |
| | Male | 7,660 (5) | 45.48 | 11,980 (5) | 49.52 | 19,640 (5) | 47.86 | 47.19 | 48.53 |
| IMD | 1 Most deprived | 2,565 (5) | 45.60 | 4,455 (5) | 52.50 | 7,020 (5) | 49.75 | 48.58 | 50.91 |
| | 2 | 2,605 (5) | 53.88 | 4,005 (6) | 55.11 | 6,610 (5) | 54.62 | 53.30 | 55.93 |
| | 3 | 2,305 (5) | 50.89 | 3,395 (6) | 55.55 | 5,700 (5) | 53.57 | 52.18 | 54.96 |
| | 4 | 1,860 (5) | 48.63 | 2,855 (5) | 52.46 | 4,715 (5) | 50.88 | 49.43 | 52.33 |
| | 5 Least deprived | 1,290 (4) | 40.46 | 2,325 (4) | 44.10 | 3,615 (4) | 42.73 | 41.34 | 44.12 |
| | Unknown | 255 (4) | 40.00 | 75 (6) | 55.15 | 330 (4) | 42.66 | 38.06 | 47.27 |
| Ethnicity | African | 60 (4) | 36.47 | 265 (4) | 41.54 | 325 (4) | 40.50 | 36.10 | 44.90 |
| | Bangladeshi or British Bangladeshi | 35 (5) | 46.98 | 155 (5) | 47.62 | 190 (5) | 47.50 | 40.75 | 54.25 |
| | British or Mixed British | 6,245 (6) | 59.01 | 10,890 (6) | 63.98 | 17,135 (6) | 62.08 | 61.15 | 63.00 |
| | Caribbean | 65 (10) | 104.84 | 205 (8) | 83.00 | 270 (9) | 87.38 | 76.96 | 97.80 |
| | Chinese | 20 (5) | 54.79 | 35 (4) | 42.42 | 55 (5) | 46.22 | 34.00 | 58.43 |
| | Indian or British Indian | 110 (6) | 63.22 | 220 (6) | 59.06 | 330 (6) | 60.38 | 53.87 | 66.90 |
| | Irish | 25 (7) | 66.67 | 80 (10) | 96.39 | 105 (9) | 87.14 | 70.47 | 103.80 |
| | Other | 55 (4) | 43.14 | 110 (4) | 39.22 | 165 (4) | 40.44 | 34.27 | 46.61 |
| | Other Asian | 70 (6) | 56.68 | 160 (5) | 47.76 | 230 (5) | 50.16 | 43.68 | 56.65 |
| | Other Black | 45 (7) | 71.43 | 195 (7) | 71.04 | 240 (7) | 71.11 | 62.11 | 80.11 |
| | Other mixed | 45 (3) | 34.48 | 105 (3) | 33.98 | 150 (3) | 34.13 | 28.67 | 39.59 |
| | Other White | 190 (4) | 37.11 | 480 (5) | 46.69 | 670 (4) | 43.51 | 40.21 | 46.80 |

| | | | | | | | | | |
|---|---|---|---|---|---|---|---|---|---|
| | Pakistani or British Pakistani | 125 (5) | 50.92 | 275 (5) | 51.35 | 400 (5) | 51.22 | 46.20 | 56.24 |
| | Unknown | 3,690 (4) | 36.74 | 3,685 (3) | 34.62 | 7,375 (4) | 35.65 | 34.84 | 36.46 |
| | White + Asian | 30 (4) | 40.27 | 60 (4) | 41.52 | 90 (4) | 41.10 | 32.61 | 49.59 |
| | White + Black African | 25 (4) | 39.06 | 65 (4) | 44.98 | 90 (4) | 43.17 | 34.25 | 52.08 |
| | White + Black Caribbean | 45 (4) | 37.19 | 125 (5) | 49.50 | 170 (5) | 45.52 | 38.67 | 52.36 |
| **Region** | East of England | 2,590 (5) | 53.05 | 595 (5) | 52.08 | 3,185 (5) | 52.86 | 51.03 | 54.70 |
| | London | 330 (4) | 37.54 | 2,935 (5) | 47.81 | 3,265 (5) | 46.53 | 44.93 | 48.12 |
| | Midlands | 2,915 (5) | 52.26 | 3,815 (7) | 66.97 | 6,730 (6) | 59.69 | 58.27 | 61.12 |
| | North East and Yorkshire | 1,855 (5) | 45.20 | 1,205 (5) | 45.62 | 3,060 (5) | 45.36 | 43.76 | 46.97 |
| | North West | 635 (3) | 31.30 | 3,340 (5) | 51.23 | 3,975 (5) | 46.50 | 45.06 | 47.95 |
| | South East | 865 (5) | 53.73 | 4,030 (5) | 48.04 | 4,895 (5) | 48.95 | 47.58 | 50.33 |
| | South West | 1,685 (5) | 47.81 | 1,190 (5) | 54.36 | 2,875 (5) | 50.32 | 48.48 | 52.16 |
| | Unknown | 10 (3) | 30.77 | 0 (0) | 0 (0) | 10 (3) | 30.77 | 11.70 | 49.84 |

**Table S9:** Counts and rates of patients with a severe mental illness currently prescribed an antipsychotic between 1st October 2021 and 31st December 2021, stratified by demographic variables

| Attribute | Category | TPP | | EMIS | | Combined | | | |
|---|---|---|---|---|---|---|---|---|---|
| | | Total (%) | Rate per 1000 | Total (%) | Rate per 1000 | Total (%) | Rate per 1000 | Lower 95% CI | Upper 95% CI |
| Total | | 80,485 (35) | 353.24 | 124,780 (38) | 381.39 | 205,265 (37) | 369.84 | 368.24 | 371.44 |
| Age group | 0-17 | 55 (22) | 215.69 | 90 (20) | 204.55 | 145 (21) | 208.63 | 174.67 | 242.59 |
| | 18-24 | 1,615 (27) | 268.27 | 2,765 (28) | 280.14 | 4,380 (28) | 275.65 | 267.48 | 283.81 |
| | 25-34 | 8,080 (28) | 278.43 | 12,735 (29) | 288.78 | 20,815 (28) | 284.67 | 280.80 | 288.54 |
| | 35-44 | 13,675 (32) | 324.24 | 21,230 (34) | 344.42 | 34,905 (34) | 336.22 | 332.70 | 339.75 |
| | 45-54 | 17,700 (37) | 371.22 | 27,760 (40) | 404.22 | 45,460 (39) | 390.70 | 387.11 | 394.29 |
| | 55-69 | 25,000 (39) | 392.83 | 39,075 (43) | 432.15 | 64,075 (42) | 415.91 | 412.69 | 419.13 |
| | 70-79 | 9,465 (37) | 370.89 | 13,965 (41) | 412.92 | 23,430 (39) | 394.84 | 389.79 | 399.90 |
| | 80+ | 4,895 (36) | 361.65 | 7,170 (39) | 393.96 | 12,065 (38) | 380.18 | 373.40 | 386.96 |
| Sex | Female | 41,725 (37) | 367.41 | 63,525 (40) | 395.47 | 105,250 (38) | 383.85 | 381.53 | 386.17 |
| | Male | 38,760 (34) | 339.15 | 61,255 (37) | 367.81 | 100,015 (36) | 356.15 | 353.94 | 358.35 |
| IMD | 1 Most deprived | 24,820 (38) | 376.57 | 41,315 (41) | 414.96 | 66,135 (40) | 399.67 | 396.62 | 402.71 |
| | 2 | 19,000 (36) | 362.53 | 31,720 (39) | 386.62 | 50,720 (38) | 377.23 | 373.94 | 380.51 |
| | 3 | 15,455 (35) | 347.30 | 21,280 (37) | 365.89 | 36,735 (36) | 357.83 | 354.17 | 361.49 |
| | 4 | 11,400 (33) | 333.58 | 16,540 (36) | 356.31 | 27,940 (35) | 346.67 | 342.61 | 350.74 |
| | 5 Least deprived | 7,975 (31) | 311.71 | 13,160 (34) | 337.00 | 21,135 (33) | 326.99 | 322.58 | 331.40 |
| | Unknown | 1,835 (35) | 348.20 | 765 (40) | 396.37 | 2,600 (36) | 361.11 | 347.23 | 374.99 |
| Ethnicity | African | 1,140 (32) | 319.33 | 3,760 (34) | 336.92 | 4,900 (33) | 332.65 | 323.34 | 341.97 |
| | Bangladeshi or British Bangladeshi | 465 (41) | 406.11 | 2,140 (46) | 459.23 | 2,605 (45) | 448.75 | 431.52 | 465.98 |
| | British or Mixed British | 44,300 (36) | 362.58 | 73,980 (40) | 401.27 | 118,280 (39) | 385.85 | 383.65 | 388.05 |
| | Caribbean | 820 (33) | 326.04 | 2,770 (35) | 352.42 | 3,590 (35) | 346.02 | 334.71 | 357.34 |
| | Chinese | 190 (36) | 355.14 | 385 (32) | 316.87 | 575 (33) | 328.57 | 301.72 | 355.43 |
| | Indian or British Indian | 1,885 (39) | 394.76 | 3,085 (41) | 408.34 | 4,970 (40) | 403.08 | 391.88 | 414.29 |
| | Irish | 520 (37) | 366.20 | 1,255 (40) | 397.15 | 1,775 (39) | 387.55 | 369.53 | 405.58 |
| | Other | 870 (33) | 327.68 | 1,495 (36) | 357.66 | 2,365 (35) | 346.01 | 332.07 | 359.96 |
| | Other Asian | 900 (35) | 350.88 | 2,180 (39) | 390.68 | 3,080 (38) | 378.15 | 364.79 | 391.50 |
| | Other Black | 395 (29) | 292.59 | 1,510 (35) | 345.93 | 1,905 (33) | 333.33 | 318.36 | 348.30 |
| | Other mixed | 345 (29) | 288.70 | 900 (33) | 327.87 | 1,245 (32) | 315.99 | 298.44 | 333.54 |

| | | | | | | | | | |
|---|---|---|---|---|---|---|---|---|---|
| | Other White | 2,705 (30) | 303.08 | 5,760 (33) | 329.43 | 8,465 (32) | 320.52 | 313.69 | 327.35 |
| | Pakistani or British Pakistani | 2,075 (40) | 403.70 | 3,465 (43) | 432.04 | 5,540 (42) | 420.97 | 409.89 | 432.06 |
| | Unknown | 22,995 (34) | 343.08 | 20,295 (34) | 340.86 | 43,290 (34) | 342.04 | 338.82 | 345.26 |
| | White + Asian | 195 (28) | 282.61 | 360 (35) | 352.94 | 555 (32) | 324.56 | 297.56 | 351.56 |
| | White + Black African | 255 (33) | 326.92 | 500 (32) | 323.62 | 755 (32) | 324.73 | 301.57 | 347.89 |
| | White + Black Caribbean | 440 (32) | 317.69 | 940 (35) | 346.22 | 1,380 (34) | 336.59 | 318.83 | 354.34 |
| **Region** | East of England | 18,220 (37) | 367.75 | 3,680 (36) | 362.03 | 21,900 (37) | 366.77 | 361.92 | 371.63 |
| | London | 6,335 (31) | 305.96 | 31,115 (35) | 348.94 | 37,450 (34) | 340.84 | 337.39 | 344.29 |
| | Midlands | 17,585 (38) | 375.27 | 19,745 (39) | 387.01 | 37,330 (38) | 381.39 | 377.52 | 385.25 |
| | North East and Yorkshire | 16,230 (37) | 367.44 | 9,445 (40) | 401.66 | 25,675 (38) | 379.33 | 374.69 | 383.97 |
| | North West | 6,580 (33) | 332.16 | 31,265 (44) | 436.42 | 37,845 (41) | 413.83 | 409.66 | 418.00 |
| | South East | 5,390 (33) | 327.06 | 22,080 (37) | 367.05 | 27,470 (36) | 358.45 | 354.21 | 362.69 |
| | South West | 10,050 (33) | 334.89 | 7,450 (35) | 346.43 | 17,500 (34) | 339.71 | 334.67 | 344.74 |
| | Unknown | 90 (33) | 327.27 | 0 (0) | 0 (0) | 90 (33) | 327.27 | 259.66 | 394.89 |

### Figures

**Figure S1:** Monthly rate of **dementia patients** issued an antipsychotic in TPP (blue line) and EMIS (red line) practices; (a) shows the rates of all antipsychotic while (b) separates out the antipsychotic medications into four common types. Grey lines represent the start of the coronavirus lockdowns in England.

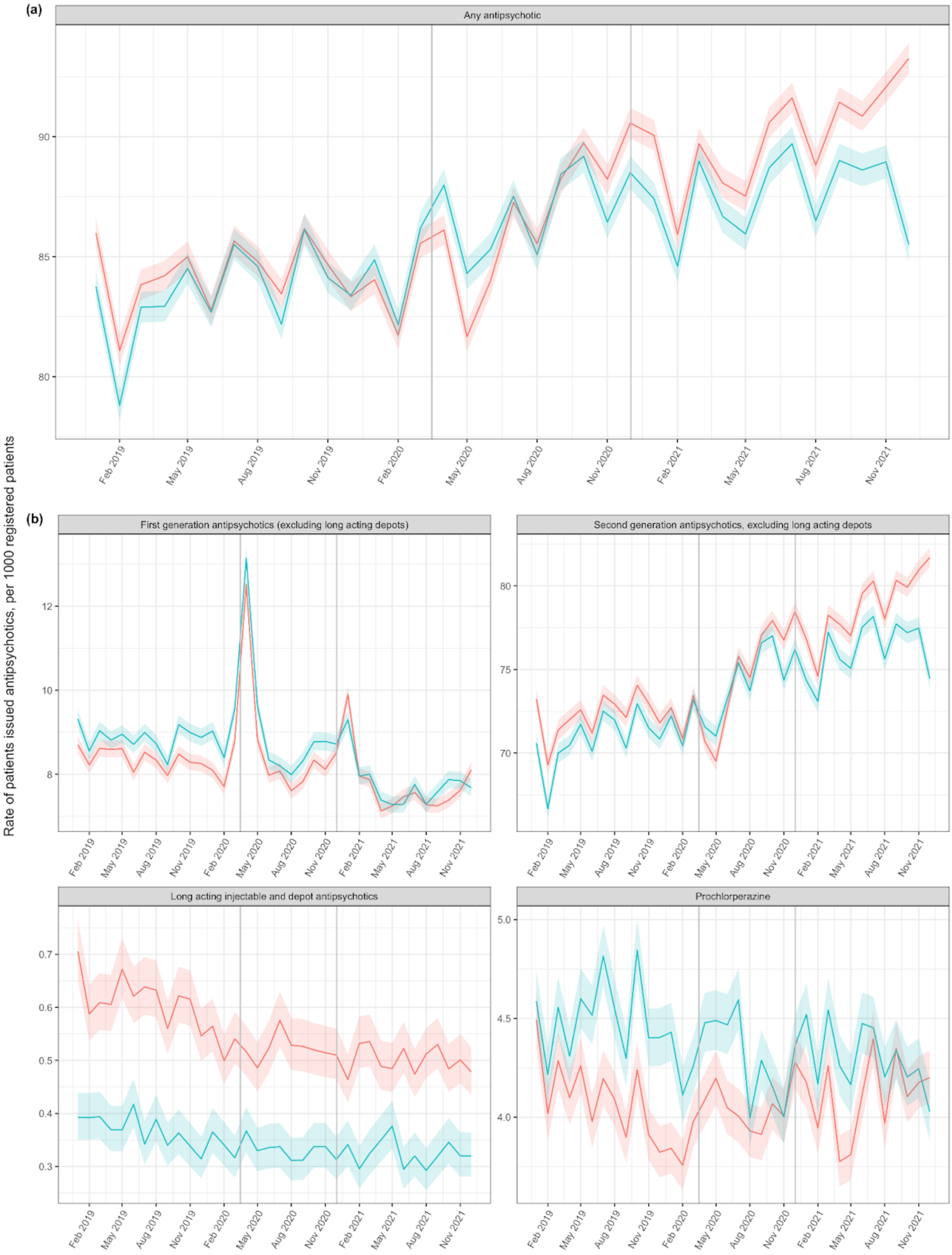

**Figure S2:** Monthly rate of **dementia patients newly issued** an antipsychotic in TPP (blue line) and EMIS (red line) practices; (a) shows the rates of all antipsychotic while (b) separates out the antipsychotic medications into four common types. Grey lines represent the start of the coronavirus lockdowns in England.

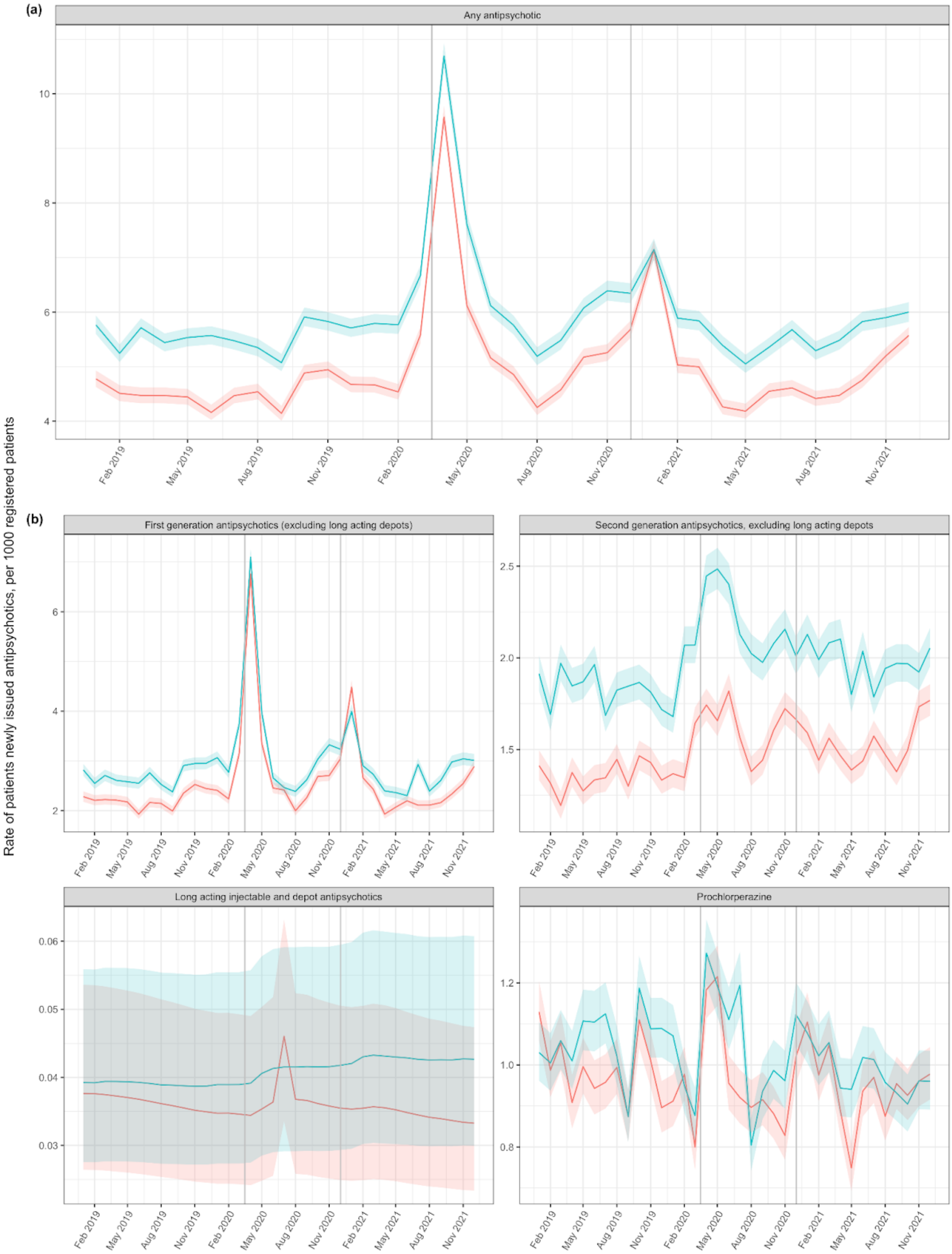

**Figure S3:** Monthly rate of **care home patients** issued an antipsychotic in TPP (blue line) and EMIS (red line) practices; (a) shows the rates of all antipsychotic while (b) separates out the antipsychotic medications into four common types. Grey lines represent the start of the coronavirus lockdowns in England.

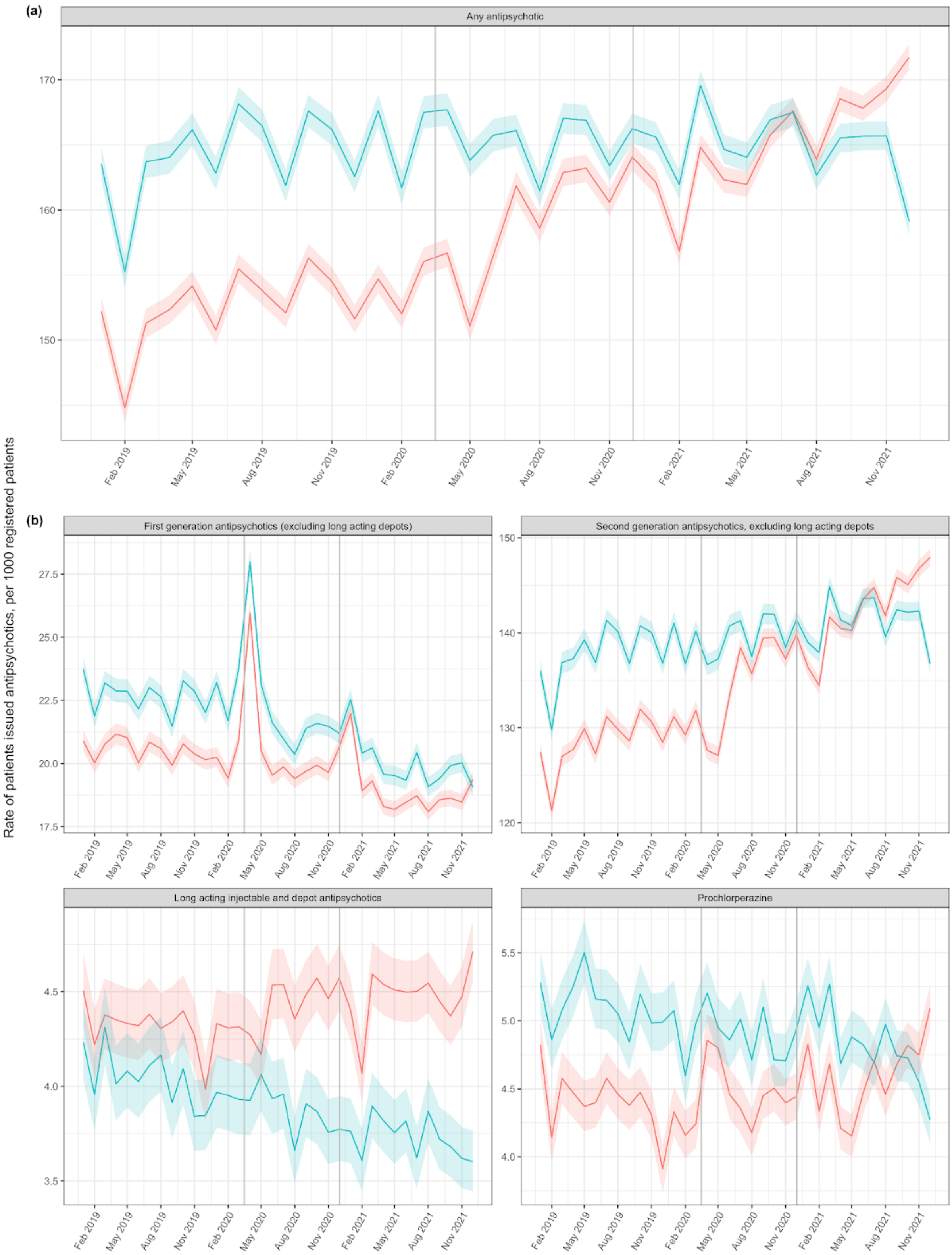

**Figure S4:** Monthly rate of **care home patients newly issued** an antipsychotic in TPP (blue line) and EMIS (red line) practices; (a) shows the rates of all antipsychotic while (b) separates out the antipsychotic medications into four common types. Grey lines represent the start of the coronavirus lockdowns in England.

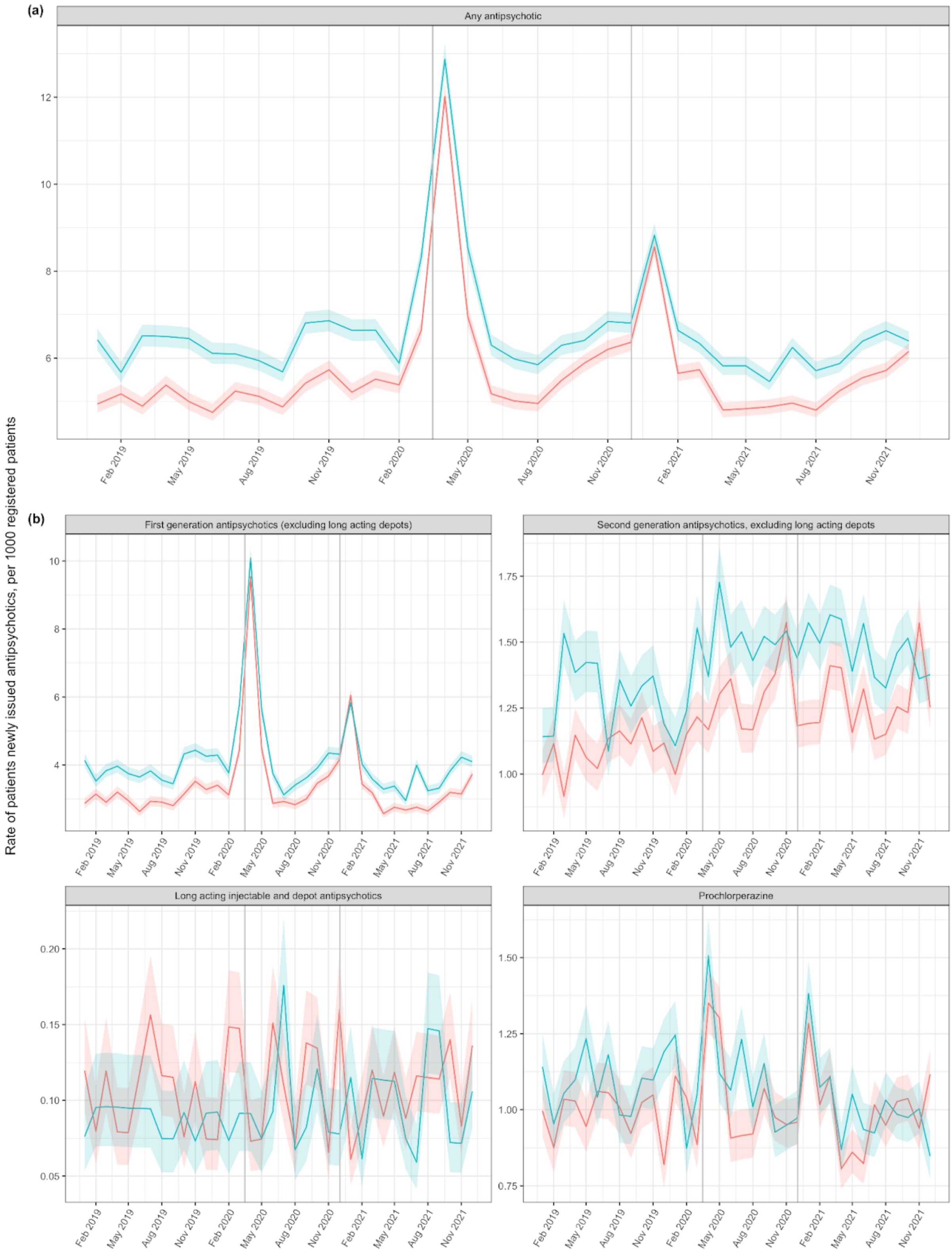

**Figure S5:** Monthly rate of **patients with a learning disability** issued an antipsychotic in TPP (blue line) and EMIS (red line) practices; (a) shows the rates of all antipsychotic while (b) separates out the antipsychotic medications into four common types. Grey lines represent the start of the coronavirus lockdowns in England.

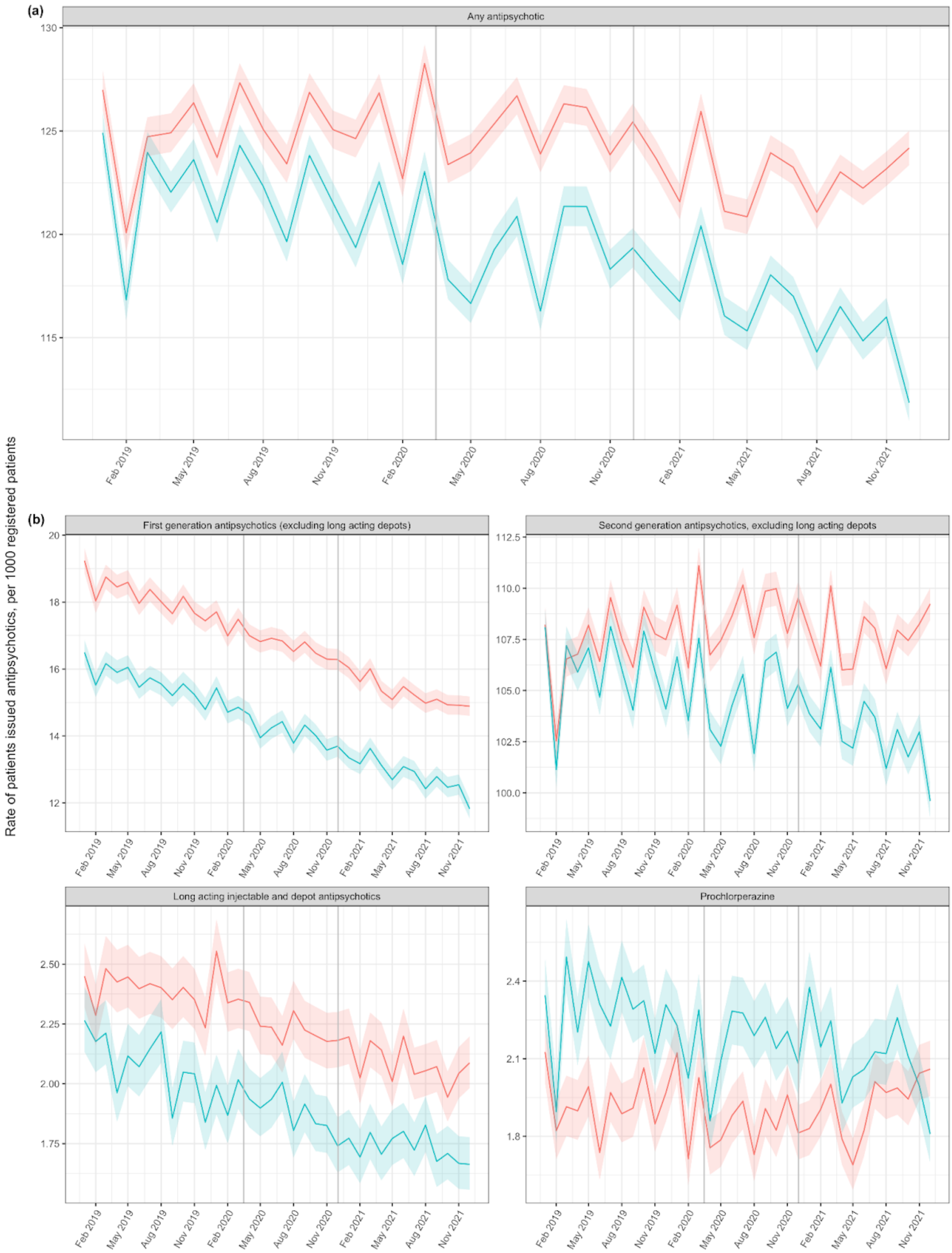

**Figure S6:** Monthly rate of **patients with a learning disability newly issued** an antipsychotic in TPP (blue line) and EMIS (red line) practices; (a) shows the rates of all antipsychotic while (b) separates out the antipsychotic medications into four common types. Grey lines represent the start of the coronavirus lockdowns in England.

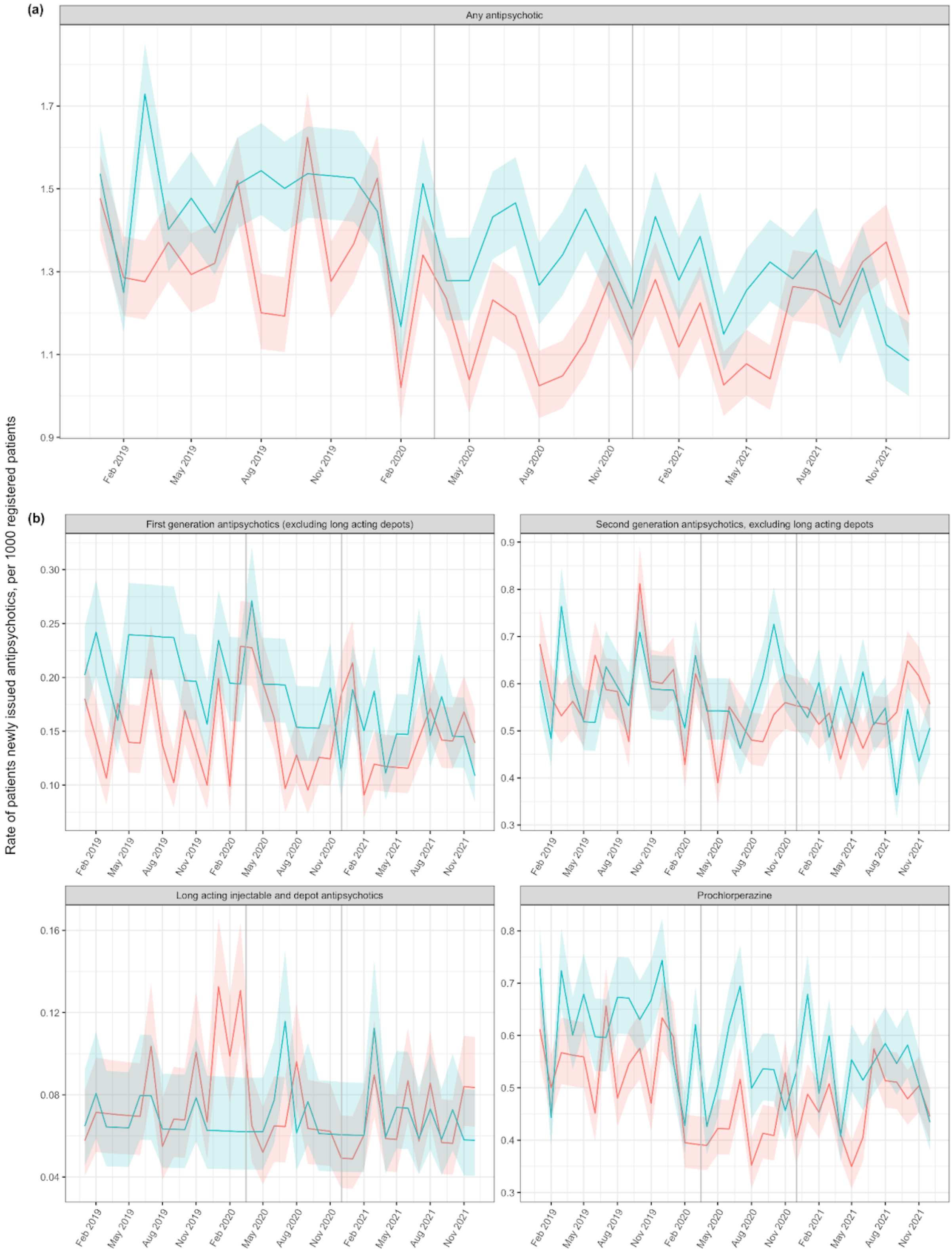

**Figure S7:** Monthly rate of **patients with autism** issued an antipsychotic in TPP (blue line) and EMIS (red line) practices; (a) shows the rates of all antipsychotic while (b) separates out the antipsychotic medications into four common types. Grey lines represent the start of the coronavirus lockdowns in England.

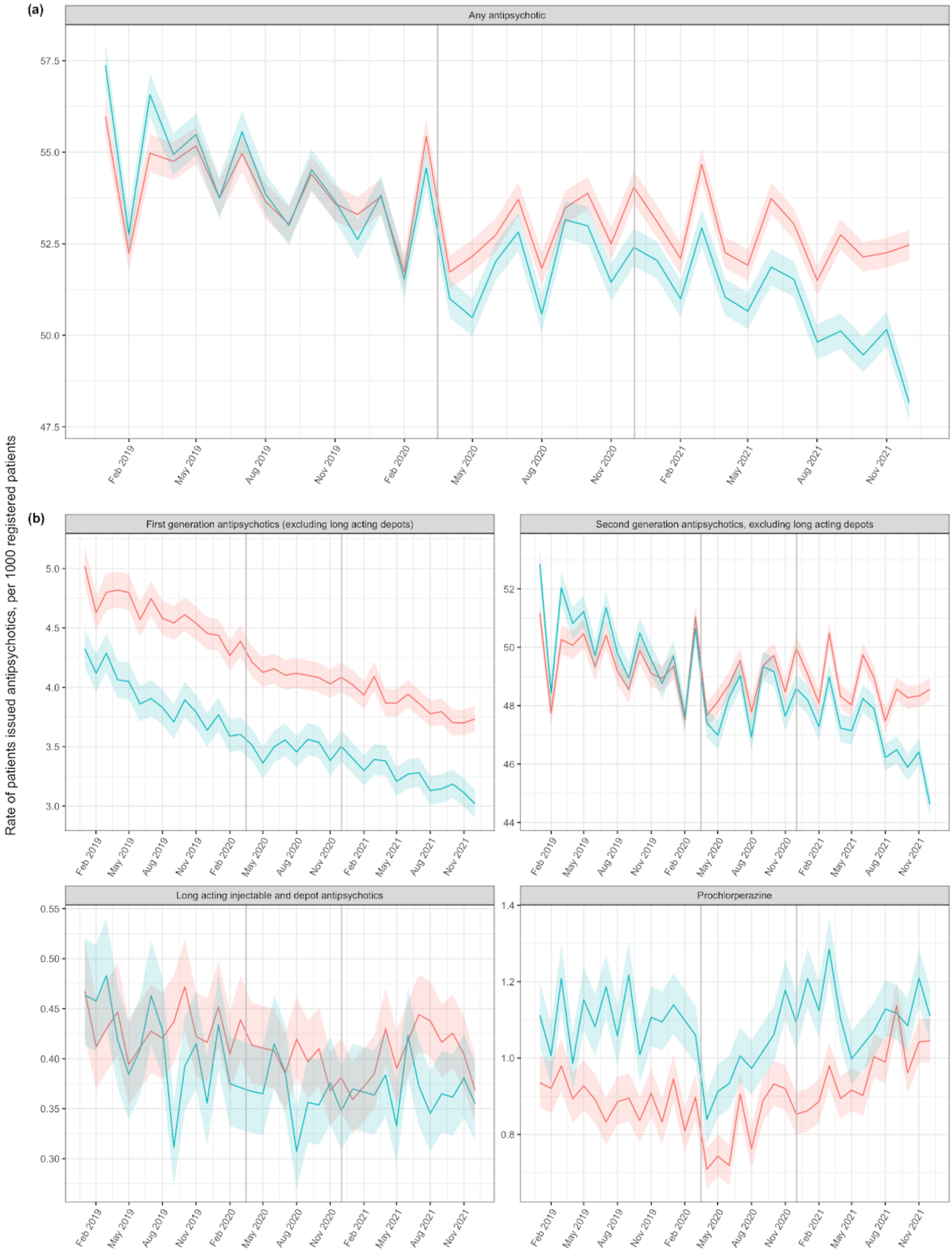

**Figure S8:** Monthly rate of **patients with autism newly issued** an antipsychotic in TPP (blue line) and EMIS (red line) practices; (a) shows the rates of all antipsychotic while (b) separates out the antipsychotic medications into four common types. Grey lines represent the start of the coronavirus lockdowns in England.

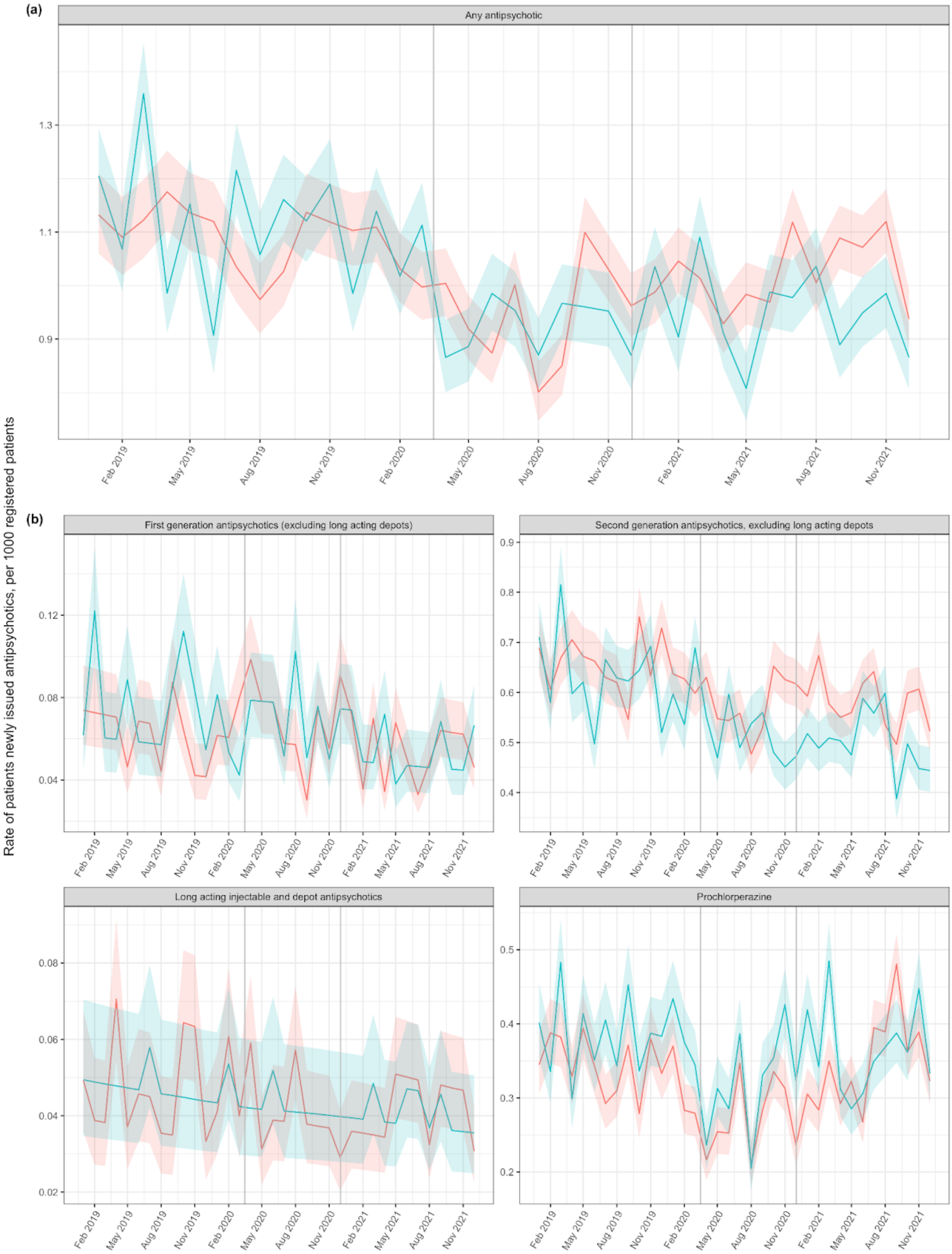

**Figure S9:** Monthly rate of **patients with a severe mental illness** issued an antipsychotic in TPP (blue line) and EMIS (red line) practices; (a) shows the rates of all antipsychotic while (b) separates out the antipsychotic medications into four common types. Grey lines represent the start of the coronavirus lockdowns in England.

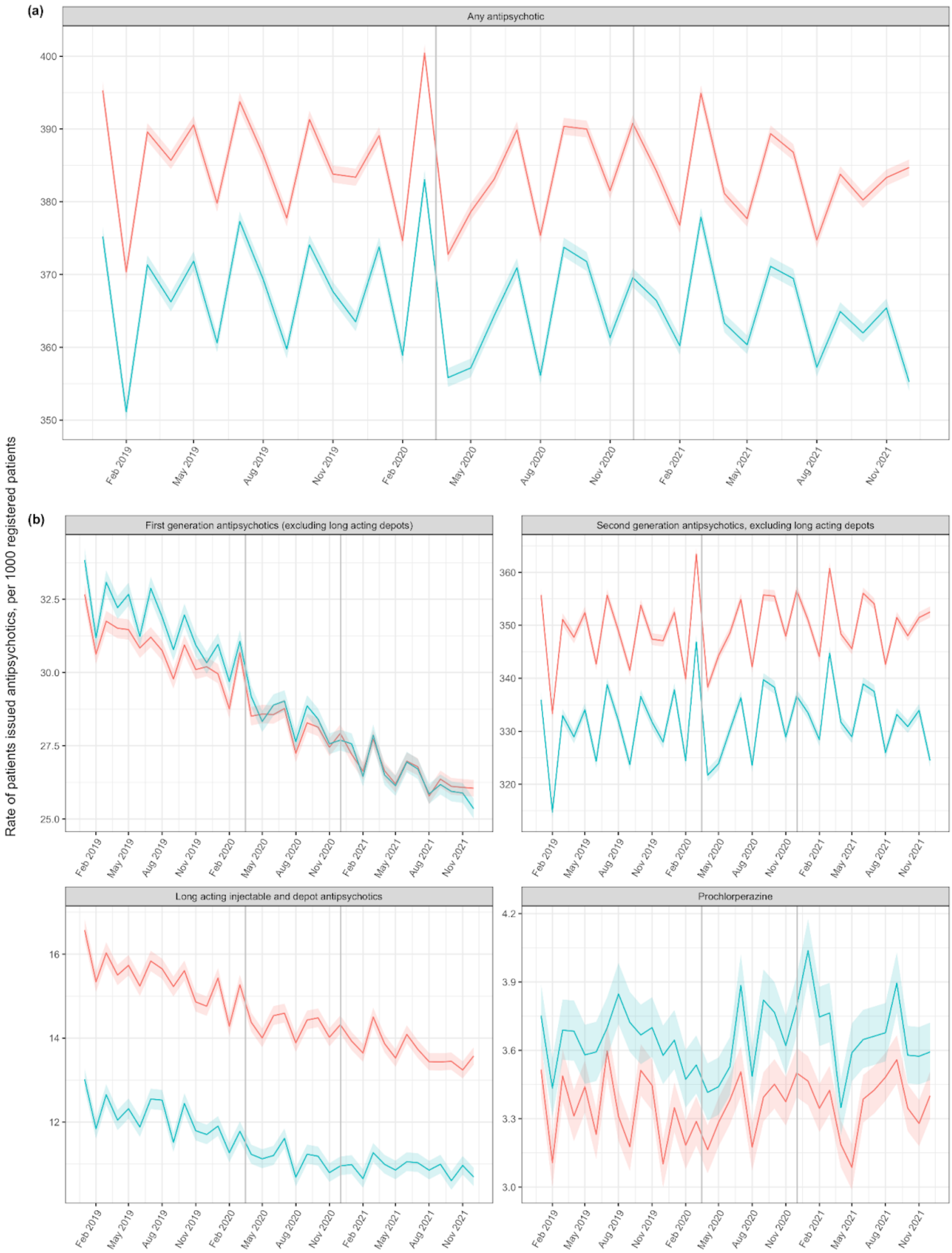

**Figure S10:** Monthly rate of **patients with a severe mental illness newly issued** an antipsychotic in TPP (blue line) and EMIS (red line) practices; (a) shows the rates of all antipsychotic while (b) separates out the antipsychotic medications into four common types. Grey lines represent the start of the coronavirus lockdowns in England.

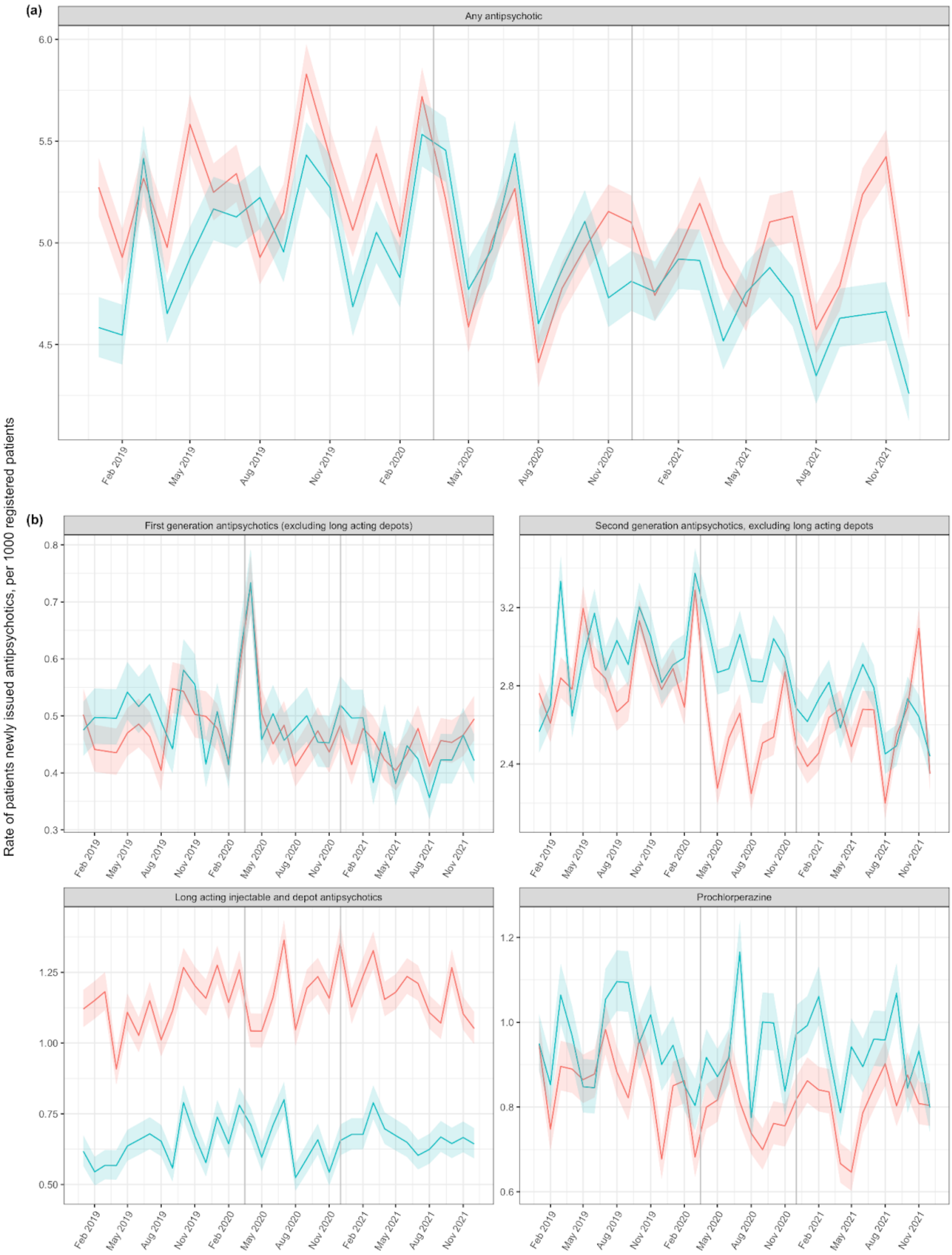
